# Disrupted Brain Hierarchical Organization in Alzheimer’s Disease Progression

**DOI:** 10.64898/2025.12.11.25342039

**Authors:** Guillermo Montaña-Valverde, Noelia Martínez-Molina, David Aquilué-Llorens, Gustavo Patow, Morten L. Kringelbach, Wolfram Hinzen, Gustavo Deco, the Alzheimer’s Disease Neuroimaging Initiative

## Abstract

Alzheimer’s disease (AD) arises from synergistic interactions between amyloid-β, tau, and neurodegeneration, yet it remains unclear how these mechanisms reshape the hierarchical organization of large-scale brain dynamics. Here, we quantified directed causal interactions across 134 participants spanning the AD continuum, comprising 46 amyloid-negative healthy controls (HC–), 36 amyloid-positive healthy controls (HC+), 31 amyloid-positive individuals with mild cognitive impairment (MCI+), and 21 amyloid-positive patients with AD dementia (AD+). Resting-state fMRI was modelled using generative effective connectivity, and hierarchical organization was assessed via trophic levels and directedness. Healthy older adults exhibited a canonical hierarchy in which sensorimotor and frontoparietal regions acted as causal sources, the default mode network (DMN) occupied an intermediate mediating position, and visual–limbic areas functioned as sinks. In contrast, AD+ individuals exhibited elevated trophic levels in the visual network and reduced levels in somatomotor, salience, control, and DMN systems. This shift was accompanied by decreased directedness, indicating a more flattened and less stratified architecture with reduced computational flexibility. MCI+ participants exhibited disruptions in somatomotor and dorsal attention networks. Compared to early-stage HC+, visual and DMN showed similar alterations while the control system returned to baseline before decreasing in AD+. Machine-learning classification distinguished all stages, including subtle differences between HC– and HC+. Hierarchical alterations were shaped by ATN biomarkers and strongly associated with cognitive decline, highlighting trophic metrics as sensitive neuroimaging biomarkers of AD progression. Together, these findings suggest that reduced hierarchical structure represents a core systems-level signature of AD, offering a promising avenue for early detection and therapeutic targeting.

## Introduction

Alzheimer’s disease (AD) is the most common cause of dementia (World Alzheimer Report, 2018). Clinically, AD presents with a devastating constellation of symptoms including memory loss and/or cognitive problems (Frisoni et al., 2010; Lam et al., 2013; Masters et al., 2015; Scheltens et al., 2021). Mild Cognitive Impairment (MCI) is the prodromal phase of AD, characterized by objective impairment in cognitive function that exceeds normal age-related changes but does not significantly affect daily activities (Petersen et al., 2018). Yet clinical diagnosis of AD based on symptoms alone is insufficient to reliably predict the presence or absence of the pathology in the brain.

Neurodegeneration, along with Amyloid-β (Aβ) deposition and pathological tau, are currently the main biomarkers for neuropathological change in the AD field (Pini et al., 2016), as defined by the ATN classification (Amyloid, Tau and Neurodegeneration; Jack et al., 2018). However, the disease can present various symptoms besides amnestic dementia, and brain changes associated with AD can occur without noticeable symptoms (Jack et al., 2018). Therefore, a better understanding of biomarkers and pathology is necessary to improve AD diagnosis and classification, allowing for the detection of early-stage neurophysiological changes and more accurate tracking of disease progression. Exploring changes in the spatiotemporal hierarchical organization of brain dynamics across the AD spectrum and its relation with ATN biomarkers could extend prior research and yield deeper insight into disease pathogenesis, ultimately supporting improved patient management and enhancing the stratification of participants in clinical trials.

Healthy brain dynamics depend on maintaining a state far from thermodynamic equilibrium, where temporal asymmetries due to irreversible processes and continuous production of entropy enable information flow, breaking the detailed balance, and establishing the “arrow of time” (Eddington, 1928; Seif et al., 2021; Perl et al., 2021; Deco et al., 2023). This temporal signature is essential for any living system, supporting information processing, adaptive behavior, and maintenance of complex computations crucial for both survival and function (Landauer, 1961; Langton, 1990; England, 2013). Previous studies revealed higher levels of nonequilibrium (i.e., higher irreversibility) during cognitively demanding tasks, implying a need for higher computational demand (Deco et al., 2022; Deco et al., 2023; Kringelbach et al., 2023). Conversely, states of reduced consciousness —such as sedation or deep sleep—showed higher reversible dynamics, suggesting constrained computational capabilities (Deco et al., 2022; García-Guzmán et al., 2023).

Growing evidence suggests that AD shifts brain dynamics toward a more equilibrium-like regime, with less complex neural activity and a decrease in entropy production (Sun et al., 2020) affecting both cognition (Chand et al., 2018) and intrinsic connectivity (Moguilner et al., 2021). Recent findings also show that temporal irreversibility negatively correlates with cognitive decline (Cruzat et al., 2023). Hierarchical organization is tightly linked to temporal asymmetry, where highly reciprocal, bidirectional interactions yield a more flattened hierarchy constraining long-range information flow, while the breaking of the detailed balance allows for a more efficient and flexible network reflected in a more vertically layered hierarchy. Despite existing evidence on nonequilibrium deviations in AD, no previous study has analysed potential differences in the brain’s hierarchical organization across the AD continuum.

The hierarchical organization of the brain has been examined from multiple complementary perspectives, each aiming to characterize the arrangement and interaction of distributed cortical and subcortical regions. Classical frameworks have described hierarchy through the topological sequence of projections (Mesulam, 1998), laminar-specific patterns of anatomical connectivity (Felleman and Van Essen, 1991), or the progression of organizational scales across structural and functional networks (Sporns, 2016). More recent approaches have shifted toward quantifying hierarchical structure through directed interactions, leveraging methods such as Granger causality (Seth et al., 2013), transfer entropy (Brovelli et al., 2015; Deco et al., 2021), and dynamic causal modelling (Friston et al., 2003; Frässle et al., 2017; Prando et al., 2020). Parallel work using functional connectivity gradients has revealed continuous cortical axes—from unimodal sensory–motor regions to transmodal association areas—through nonlinear dimensionality reduction (Margulies et al., 2016). Moreover, gradients of structure–function coupling have demonstrated a transition from tightly coupled sensory–motor regions to markedly decoupled higher-order cortical territories (Preti and De Ville, 2019). Although these gradient-based frameworks provide valuable insights into functional similarity (Hu et al., 2022; He et al., 2023) and the alignment between structural and functional architecture (Sun et al., 2024), they remain correlation-based and therefore undirected. As such, they are limited in their capacity to capture asymmetries in information flow. In contrast, the present study adopts a mechanistic framework grounded in causal interactions, allowing for a more comprehensive characterization of hierarchical brain organization.

Here, we operationalize hierarchy by applying a measure inspired by ecological network theory, where trophic levels describe the relative “height’’ of nodes within a directed graph (Levine, 1980). In our case, trophic levels are computed for each brain region using the Generative Effective Connectivity (GEC) estimated via whole-brain modelling for each participant (Kringelbach et al., 2023; Fig. 1), which corresponds to asymmetric weights assigned to the existing anatomical connections. This approach extends the classic notion of effective connectivity (Friston et al., 2003), with the distinction that GEC is *generative*: the whole-brain model iteratively adjusts the strengths of anatomical pathways—effectively the conductive values of each fibre—to best reproduce the empirical resting-state dynamics. This framework enables a principled examination of directed network architectures and their functional implications (MacKay et al., 2020), and has recently been extended to characterise pharmacologically induced rebalancing of large-scale brain dynamics (Deco et al., 2024). We adopt a definition of hierarchy as the vertical organization of regions that best captures the directionality and influence embedded in their interactions, derived by minimizing a hierarchy energy function that incorporates both edge weights and directions (Carmel et al., 2002). The resulting *trophic levels* specify the hierarchical position of each region, while the *trophic directedness* quantifies the coherence of the overall hierarchy—namely, the extent to which directed connections align along a consistent global ordering. Regions with high trophic levels act as sources, driving causal output; those with intermediate levels function as mediators, balancing incoming and outgoing influences; and those with low trophic levels act as sinks, where information converges. This framework thus provides both regional and global markers of hierarchical structure, grounded in a mechanistic model of directed causal interactions.

**Figure 1.**
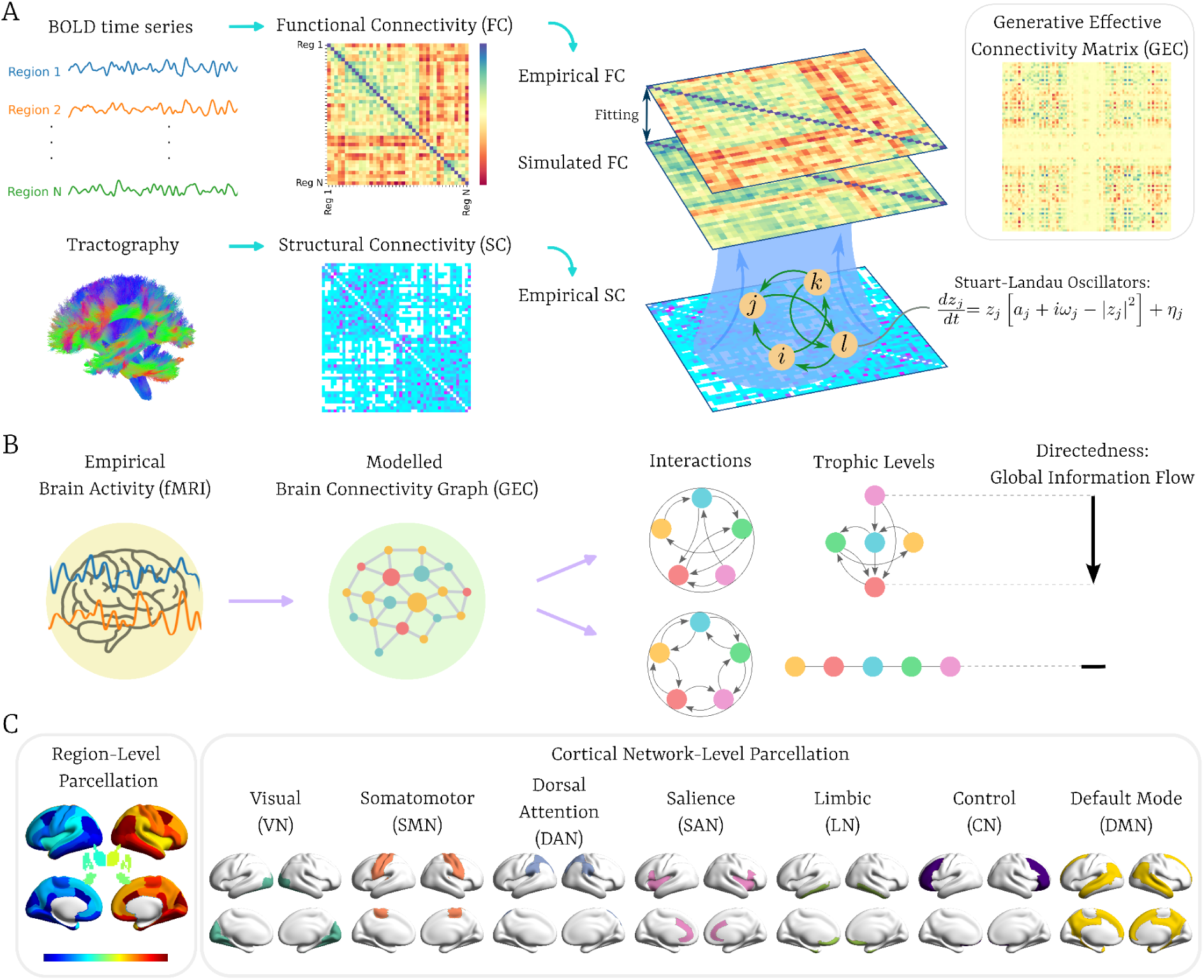
Pipeline for assessing hierarchy in the brain using trophic levels and directedness. **(A)** Time series were extracted from denoised, preprocessed Blood Oxygenation Level Dependent (BOLD) signals measured by functional MRI. These were used to compute the functional connectivity (FC), which, together with structural connectivity (SC) derived from diffusion weighted imaging (DWI) tractography, was employed to compute anatomically constrained generative effective connectivity (GEC). **(B)** GEC was then represented as a directed graph to compute the brain’s hierarchical organization at the level of each node, i.e., brain region or network. **(C)** The Desikan-Killiani brain atlas, consisting of 62 regions together and 18 subcortical areas, was used for brain parcellation. Additionally, these regions were classified into 7 distinct functional networks based on the Schaefer parcellation by matching each region to its corresponding network through overlapping (see Methods).

In this article, the main aim is to study how the brain’s hierarchical organization is reconfigured in the AD continuum using cross-sectional data from the ADNI dataset, including resting-state fMRI, PET imaging, and a battery of cognitive tests. Our AD staging was based on cognitive performance and amyloid-beta status, consistent with the amyloid cascade framework (Hardy & Higgins, 1992), and goes in line with recent studies on fluid biomarkers (Montoliu-Gaya et al., 2025) and rsfMRI alterations (Roemer-Cassiano et al., 2025).

We hypothesized that: (i) the brain’s hierarchical organization would be significantly altered in AD+ individuals compared with healthy controls and MCI+ participants, and that these alterations would be reflected in whole-brain directedness; (ii) a machine-learning classifier would reliably discriminate disease stages based on trophic levels and directedness, indicating that these hierarchical features contain intrinsic diagnostic information relevant to AD progression; and (iii) hierarchically altered brain regions would show significant associations with ATN biomarkers and cognitive performance, thereby linking hierarchical organization to disease severity.

## Results

To address our main research question, we quantified hierarchical reconfiguration across the AD continuum. As illustrated in Fig. 1, we estimated the GEC for each participant and derived global directedness and regional trophic levels to characterize hierarchical organization at different AD stages. This cross-sectional design enabled us to examine how hierarchical architecture varies as a function of Aβ status and cognitive impairment severity, thereby linking alterations in directed interactions to the progression of AD pathology.

### Characterization of Trophic Levels in Amyloid-Negative Healthy Elderly Controls

To establish a normative reference before assessing hierarchical disruption across the AD continuum, we first quantified trophic levels in a healthy elderly cohort (HC–). Partitioning the z-scored distribution into three equal ranges (Fig. 2a) revealed a robust stratification of the cortex into source, mediator and sink regimes (Fig. 2b). Source regions were located primarily in sensorimotor and fronto-parietal cortices, including the precentral and postcentral gyri, paracentral lobule, inferior frontal subdivisions, parietal regions, dorsomedial prefrontal cortex, cingulate cortices, insula and putamen. This pattern indicates that, in the resting state, motor-related circuits and basal ganglia structures occupy dominant positions within the causal flow architecture.

**Figure 2.**
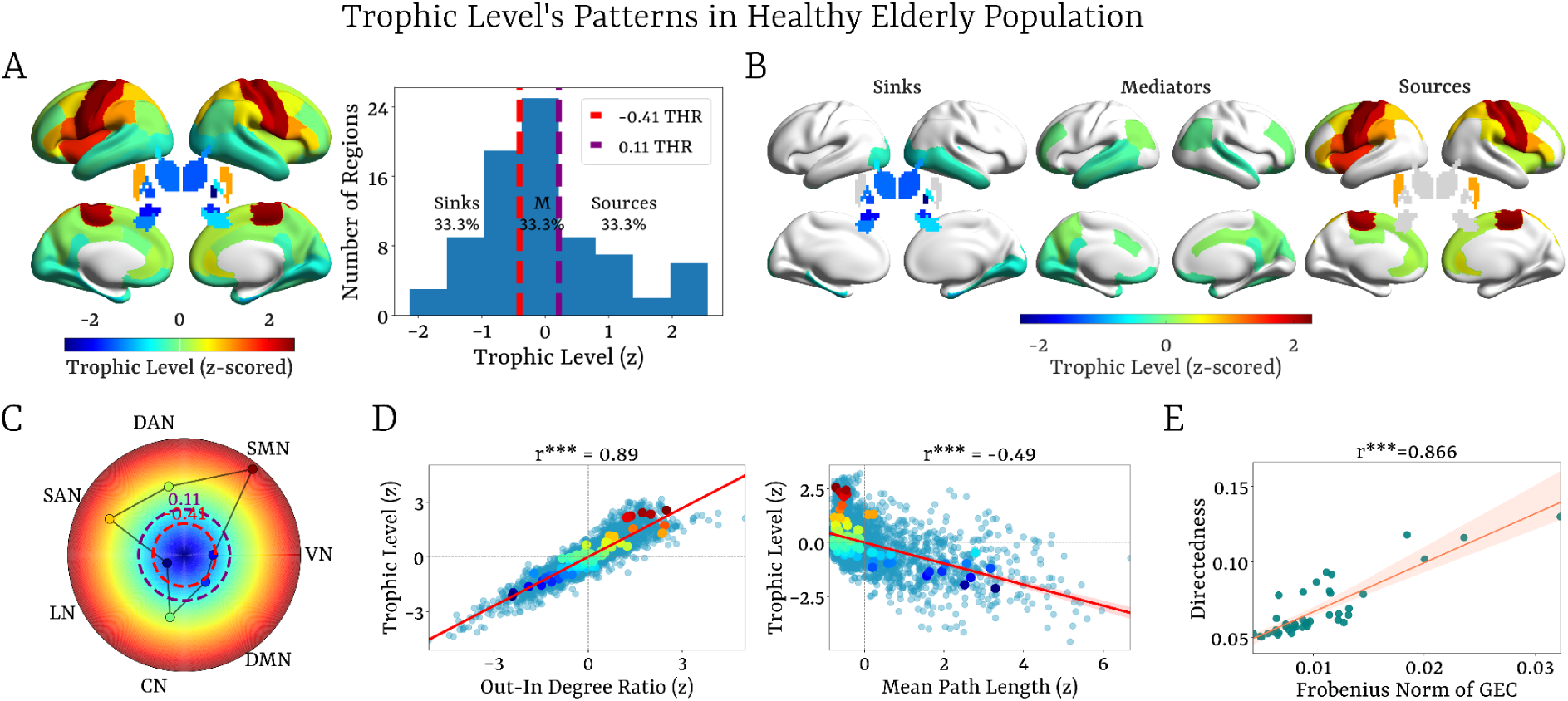
Trophic Level’s Patterns in the Healthy Elderly Population (HC−). **(A)** Z-scored trophic levels across the cortex and subcortex, and their distributions were divided into three equal ranges, revealing the classification of brain regions into sinks, mediators, and sources. **(B)** Brain renderings displaying the three identified trophic level regimes (sinks, mediators, and sources) based on z-scored trophic levels. **(C)** Network-level averaged trophic levels with corresponding thresholds for the distribution of trophic levels across the regions. **(D)** Correlations between z-scored trophic levels with the strongest and significant network properties, including the local imbalance/out-in-degree ratio (r=0.89, p<0.001) and mean path length (r=-0.49, p<0.001), showing the influence of causal imbalance and communication efficiency on trophic levels. **(E)** Correlation between directedness and the Frobenius norm of each individual’s GEC (r=0.866, p<0.001), implying that verticality in the hierarchical organization is strongly linked to the asymmetry of causal interactions.

Mediator regions encompassed the rostral middle frontal, orbitofrontal, precuneus, cuneus, and cingulate cortices, angular gyrus, and temporal lobe areas. Sink regions were predominantly observed in occipital, fusiform, entorhinal, and parahippocampal cortices, and in subcortical areas including the thalamus, hippocampus, amygdala, caudate, and globus pallidus. Together, these regimes delineate a large-scale gradient extending from sensorimotor, salience, dorsal attention and control networks acting as propagators, through integrative DMN territories, to visual and limbic networks where information converges (Fig. 2c). Lower trophic levels in visual cortices at rest likely reflect stronger incoming influence, consistent with predictive processing in the absence of external stimulation (Richter et al., 2024). This interpretation is supported by increased trophic levels during task performance (Supplementary Fig. 2) and by replication of this hierarchical scaffold in an independent HCP-Aging cohort (Supplementary Fig. 3).

Regression analyses showed that trophic levels were dominated by local imbalance/out-in degree ratio (r = 0.89, p < 0.001), followed by mean path length (r = –0.49, p < 0.001; Fig. 2d). Both clustering coefficient and betweenness centrality showed significant but weak positive correlations (r < 0.05). Overall, these variables–local imbalance/out-in degree ratio, mean path length, clustering coefficient, and betweenness centrality–explained a large proportion of variance (R² = 0.85), indicating that trophic levels principally capture asymmetries between outgoing and incoming influences, while also reflecting broader communication efficiency within the effective connectivity network.

Collectively, the gradient of trophic levels outlines the functional scaffold of directed interactions, positioning each region within a feedforward–feedback architecture constrained by anatomical connectivity. This organization does not imply a unidimensional ranking of regions; rather, functional relevance emerges from the intermediary mediator layer that balances inflow and outflow and coordinates distributed processing. Notably, the DMN mapped closely onto this mediator layer, highlighting its central role in orchestrating bidirectional information flow between source and sink systems.

The strong correlation between directedness and the Frobenius norm of each individual’s GEC (r = 0.866, p< 0.001; Fig. 2e) suggests that the verticality of the hierarchical organization of a directed network is closely linked to its asymmetric connections; in other words, to the interactions of causal flow across brain regions. This finding underscores the central role of causal imbalance in shaping the brain’s hierarchical architecture, where bidirectional, recurrent interactions limit the system’s capacity for complex computations, while asymmetric interactions enable directed information flow, supporting complex temporal dynamics.

### Hierarchical Reconfiguration across the AD continuum

Group-level analyses at both regional and network scales revealed marked differences in hierarchical brain organization in AD+ participants compared with MCI+, HC+, and HC– groups (Fig. 3a,b). Relative to HC–, AD+ individuals showed elevated trophic levels in the visual network (VN), including the cuneus, pericalcarine, lingual, fusiform, and occipital regions, along with the bilateral thalamus and amygdala. In contrast, reduced trophic levels were observed in the somatomotor (SMN), salience (SAN), control (CN), and default mode (DMN) networks, including the left precentral and postcentral cortices, bilateral middle frontal areas (both rostral and caudal), bilateral cingulate cortices (rostral and isthmus), right posterior cingulate and precuneus, left insula, left middle temporal gyrus, left inferior frontal gyrus, bilateral angular gyrus, and right hippocampus.

**Figure 3.**
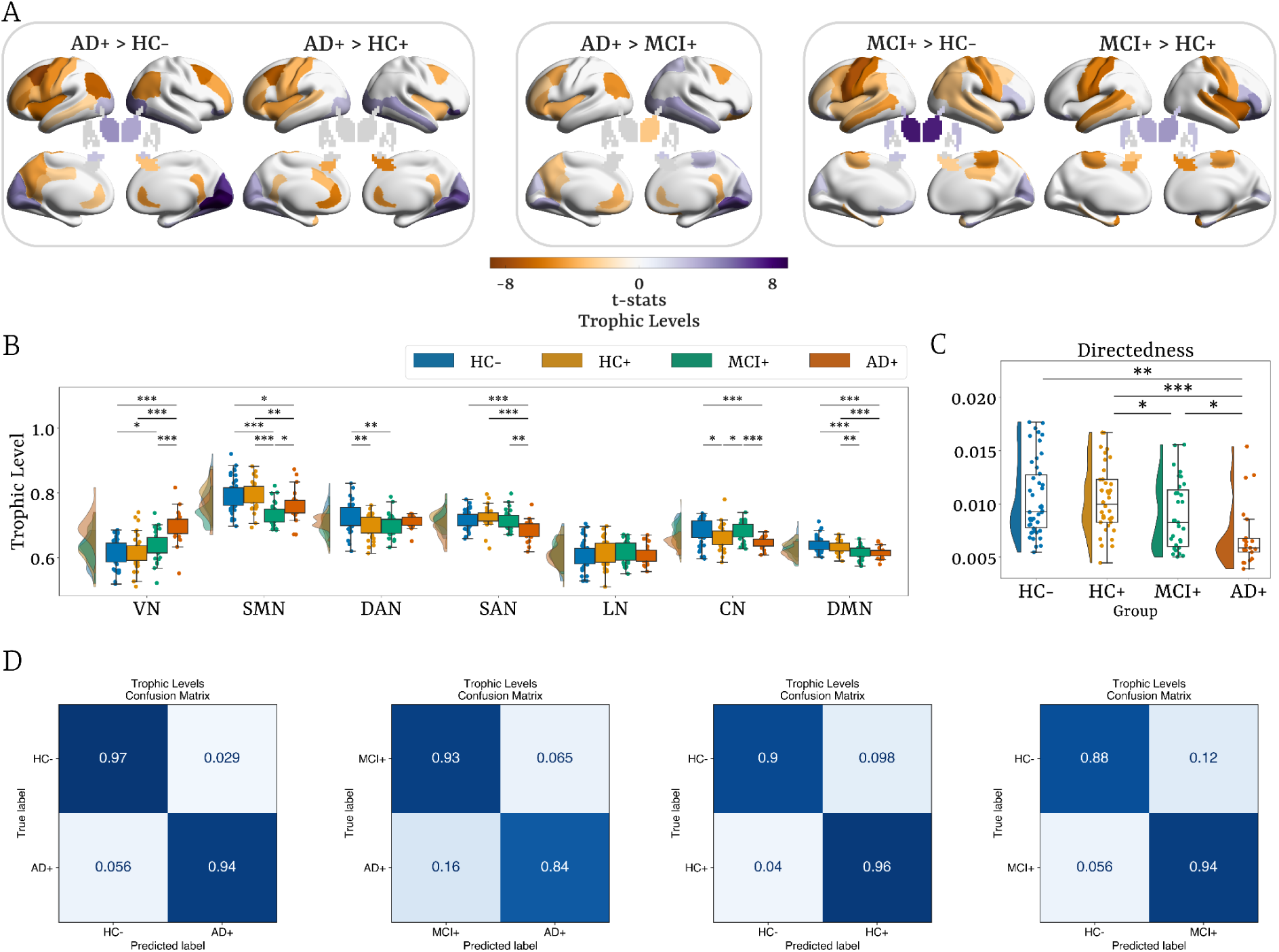
The brain’s hierarchical reorganization in the progression of AD. **(A)** Group comparison at the regional level. **(B)** Group comparisons at the network level. **(C)** Directedness comparisons across groups. **(D)** Confusion matrices on trophic levels’ machine learning-based classification performance for each pair of groups, with rows indicating true categories and columns representing predicted categories. Group comparisons were performed using non-parametric permutation tests (10,000 permutations), controlling for age, gender, and education as covariates, and FDR-corrected. HC−: amyloid-negative healthy controls, HC+: amyloid-positive healthy controls, MCI+: amyloid-positive with mild cognitive impairment (MCI+), AD+: amyloid-positive with Alzheimer’s disease. VN: visual network, SMN: somatomotor network, DAN: dorsal attention network, SAN: salience network, LN: limbic network, CN: control network, DMN: default mode network. Here and below, asterisks denote significant results after FDR correction*: * P<0.05*, *\*\* P<0.01*, \*\**\* P<0.001*.

Most alterations remained significant when comparing AD+ with HC+, except for the bilateral thalamus and amygdala, left precuneus and posterior cingulate, bilateral angular gyrus, and the right rostral middle frontal area. Additionally, decreases were observed in the left caudal anterior cingulate, right insula, and left hippocampus, and an increase in the right middle temporal gyrus. The AD+ group consistently exhibited increased trophic levels in VN and decreased levels in SMN, SAN, CN, and DMN. Comparisons with MCI+ yielded fewer significant differences, as expected with advancing pathology: increases persisted in visual regions (cuneus, pericalcarine, lingual, fusiform, and occipital) and right middle temporal gyrus, while decreases remained in the bilateral rostral anterior and isthmus cingulates, left precuneus, frontal, and angular regions, in addition to a reduction in the right thalamus.

MCI+ participants also showed evidence of hierarchical reconfiguration. Compared to both HC– and HC+, they exhibited reduced trophic levels in bilateral precentral, postcentral, and paracentral cortices, and bilateral parahippocampus, with an increase in occipital areas and the bilateral thalamus. Relative to HC–, additional reductions appeared in the right posterior cingulate cortex, bilateral superior and middle temporal gyri, left insula, and left hippocampus, along with an increase in the right inferior frontal gyrus; compared to HC+, reductions were evident in the right insula, bilateral hippocampus, and right amygdala. At the network level, these effects manifested as elevated trophic levels in VN (vs. HC–) and decreased levels in SMN, dorsal attention (DAN), and DMN (vs. both control groups). Notably, trophic levels in CN showed an early decrease in HC+, returned to baseline in MCI+, before falling again in AD+.

These spatially-specific changes were accompanied by a significant decrease in whole-brain directedness in AD+ compared with HC–, HC+, and MCI+ groups (Fig. 3c). Reduced directedness reflects diminished separation between trophic levels across regions, indicating a shift toward a less vertically structured, more flattened, and more homogeneous hierarchy—an effect present in MCI+ compared to HC+, but less pronounced. This reconfiguration points to reduced global directionality of information flow, with AD+ brain connectivity becoming more bidirectional within a less efficient and less flexible system, characterized by a weakened top-down control and enhanced bottom-up drive.

### Machine Learning-Based Classification of AD Stage

We used machine learning to establish the significant hierarchical reorganization across the different AD stages (Figure 3D). For classification, we used the regional and network-level trophic levels as well as directness as input features. We trained a logistic regression model with a leave-one-out cross-validation procedure and a grid search with three hyperparameters: number of features (NF), regularization, and penalty. For each NF, the Minimum Redundancy Maximum Relevance (mRMR) algorithm was applied to mitigate redundancy among predictors. To reduce dependence on a single data split and obtain more robust performance estimates, we performed N = 20 Monte Carlo iterations of the full model training and evaluation pipeline (see Methods).

We found that the highest classification accuracy was obtained for classifying the AD+ from the HC– group with 0.96 ± 0.04 (mean ± s.d.) accuracy. In comparison, when classifying the MCI+ group against AD+, the accuracy was 88.5 ± 0.04, remaining a strong but slightly lower performance relative to AD+ vs. HC–. Importantly, we were able to classify the HC+ preclinical stage from HC– with an accuracy of 0.93 ± 0.06. In addition, we also carried out the classification for HC– vs. MCI+, which showed an accuracy of 0.91 ± 0.05. The ROC (Receiver Operating Characteristic) endorsed a similarly high mean accuracy (Supplementary Figure 4, Table 3). All classification results were statistically significant. For all classification metrics, see Table 3.

Across classification analyses, SHAP values revealed a graded shift in the trophic level’s features contributing most strongly to discrimination (Supplementary Figure 4). In the HC– vs HC+ comparison, the highest SHAP values were confined to subcortical nodes–including the globus pallidus externus, nucleus accumbens, and amygdala–indicating early hierarchical disruptions preceding cortical involvement. Distinguishing HC– from MCI+ recruited additional thalamic, insular, and fronto-striatal regions, consistent with broader propagation of network-level alterations during prodromal impairment. By contrast, classifications involving AD+ showed the most extensive set of influential features, with both cortical (cingulate, parietal, occipital, and orbitofrontal) and subcortical regions contributing prominently. Together, these results indicate that trophic level reorganization becomes increasingly widespread with disease progression, transitioning from circumscribed subcortical alterations in preclinical stages to a more diffuse cortical–subcortical disruption in symptomatic AD.

### How does ATN Shape the Brain’s Hierarchical Organization?

Mixed-effects ridge regression revealed network-level relationships between standardized ATN biomarker and trophic levels. For Aβ, higher deposition was related to higher trophic levels in VN (β_A_ = 0.035, p<0.001), while negative associations were observed in the SMN (β_A_ = −0.018, p<0.001), SAN (β_A_ = −0.007, p<0.05), and DMN (β_A_ = −0.009, p<0.001)(Figure 4A). At the regional level, Aβ-related increases in trophic levels were localized in occipital areas, the left hippocampus, and bilateral thalamus, whereas decreases were observed in the left insula, bilateral precentral, postcentral, and paracentral, bilateral rostral and caudal middle frontal cortices, left middle temporal gyrus, left posterior cingulate, and right rostral anterior cingulate.

**Figure 4.**
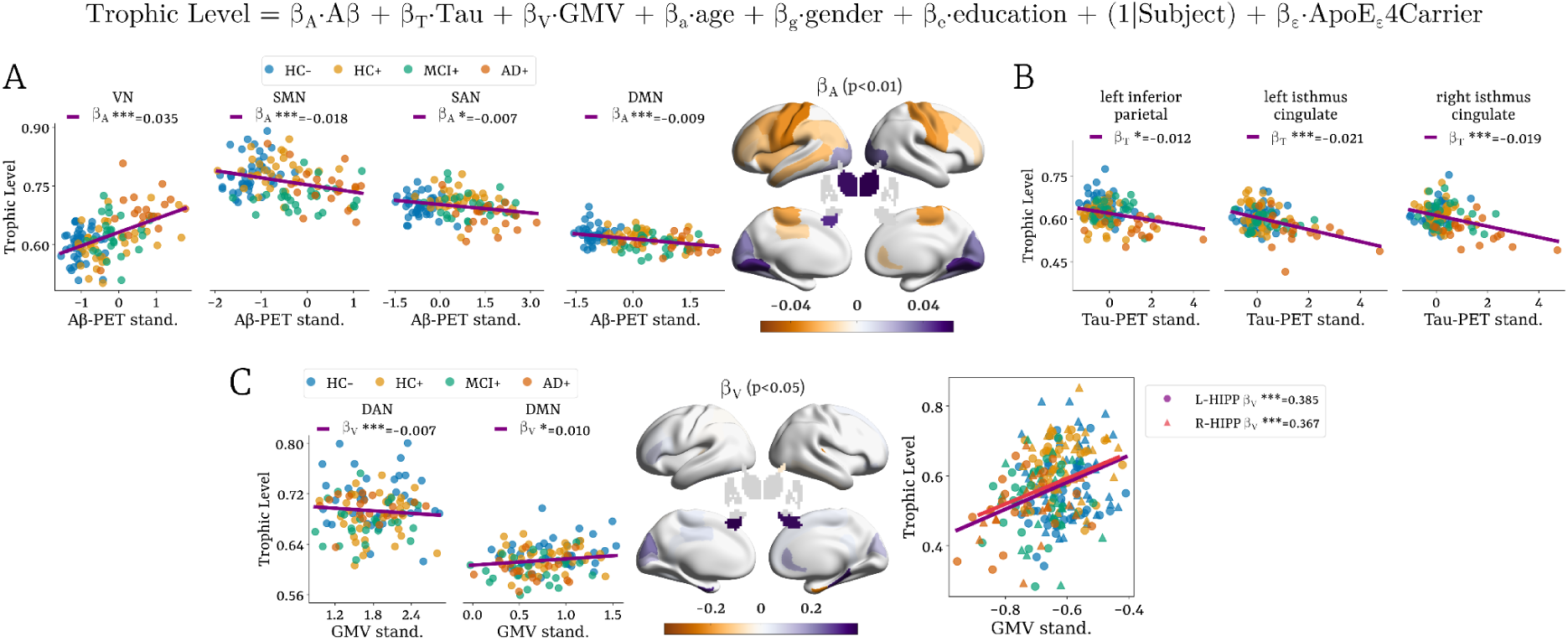
Associations between trophic levels and ATN biomarkers using mixed-effects ridge regression. **(A)** Aβ deposition was positively associated with trophic levels in the VN and negatively in SMN, SAN, and DMN. **(B)** Tau accumulation showed a negative association with trophic levels in the left inferior parietal and the bilateral isthmus. **(C)** Reduced grey matter volume (GMV) was related to higher trophic levels in the DAN and lower in the SAN and DMN, with the strongest effects in the bilateral hippocampus. All results were adjusted for covariates (age, gender, education, APOE carrier status) and a random intercept for subject was modeled to account for within-subject variability, and validated using permutation testing. VN: visual network, SMN: somatomotor network, DAN: dorsal attention network, SAN: salience network, CN: control network, DMN: default mode network.

Lower tau accumulation in the left inferior parietal (β_T_ = −0.012, p<0.05), and bilateral isthmus cingulate (left: β_T_ = −0.021, p<0.001; right: β_T_ = −0.019, p<0.001) was related to lower trophic levels (Figure 4B). Lower GMV–reflecting atrophy–was significantly related to higher trophic levels in the DAN (β_V_ = −0.007, p<0.001) and to lower trophic levels in the DMN (β_V_ = 0.010, p < 0.05). At the regional level, the strongest GMV-trophic level relationships were found in the bilateral hippocampus, where greater atrophy corresponded to lower trophic levels (Figure 4C).

### Associations Between Trophic Levels and Cognitive Decline

Multiple linear regression analyses (FDR-corrected) revealed robust associations between hierarchical organization and cognitive decline across domains. Greater impairment on ADAS-Cog-13 was linked to higher trophic levels in VN (β_TL_ = 79.6, p < 0.001) and lower trophic levels in the SMN (β_TL_ = –41.6, p < 0.05), SAN (β_TL_ = –113.2, p < 0.001), CN (β_TL_ = –61.9, p < 0.05), and DMN (β_TL_ = –131.1, p < 0.001). Similar gradients were observed for CDR-SB, where worse clinical severity was associated with elevated trophic levels in VN (β_TL_ = 13.3, p < 0.001), and reduced levels in SAN (β_TL_ = –17.4, p < 0.01) and DMN (β_TL_ = –22.4, p < 0.01). For MoCA, lower scores were associated with higher trophic levels in VN (β_TL_ = –35.9, p < 0.001), and lower trophic levels in SMN (β_TL_ = 17.8, p < 0.05), SAN (β_TL_ = 45.0, p < 0.01), and DMN (β_TL_ = 55.8, p < 0.01). MMSE followed the same pattern, with greater decline corresponding to higher trophic levels in VN (β_TL_ = –20.0, p < 0.001), and reduced levels in SAN (β_TL_ = 29.7, p < 0.001), and DMN (β_TL_ = 32.4, p < 0.01).

At the regional level, cognitive deterioration across all four assessments (ADAS-Cog-13, CDR-SB, MoCA, and MMSE) was consistently associated with lower trophic levels in the bilateral precentral, postcentral, and paracentral cortices, left rostral and caudal middle and inferior frontal regions, left angular gyrus, left insula, and bilateral hippocampus. Conversely, higher trophic levels with worsening cognition were observed in occipital areas—including cuneus, pericalcarine, lingual, and fusiform cortices—as well as in the bilateral thalamus. Lower trophic levels throughout the cingulate cortex (rostral, caudal, and isthmus subdivisions) were also strongly associated with poorer performance across cognitive measures.

## Discussion

Cognitive function depends on the brain’s ability to coordinate neural activity from local circuits to large-scale networks (Tononi et al., 1994; Sporns, 2000), supported by a flexible hierarchical architecture that adapts to changing cognitive demands (Cole et al., 2013; Deco et al., 2015). In this study, we investigated how AD reshapes the hierarchical organization of large-scale brain dynamics across the disease continuum. Directly addressing our central research question, we show that hierarchical measures derived from GEC—trophic levels and directedness—capture robust and stage-dependent alterations in the brain’s hierarchical architecture. Our results revealed hierarchical flattening in AD+, characterized by an increase in trophic levels of sink regions–specifically visual and thalamic areas–and a reduction in source and mediator nodes, including motor, salience, and higher-order DMN regions. These alterations were tightly linked to ATN biomarkers and cognitive performance, underscoring their pathologicall relevance. More broadly, the framework introduced here provides a mechanistic and principled approach for quantifying hierarchical reorganization in neurodegenerative disease, complementing prior work on structural and functional network disruption in AD. Future longitudinal studies with larger samples will be essential to determine whether hierarchical alterations precede clinical symptoms and whether they contribute to early disease detection, prognosis, or treatment monitoring.

### Hierarchical organization in healthy controls

We first established the hierarchical scaffold in amyloid-negative healthy controls. The resulting structure comprised three distinct units—sources, mediators and sinks—corresponding to sensorimotor, frontoparietal, dorsal attention and salience regions (sources), default mode network (DMN) regions (mediators), and visual and limbic areas (sinks) (Fig. 2a–c). This organization reflects the balance between feedforward and feedback processes at rest in healthy ageing.

Trophic levels were driven by both local causal imbalance and global topological properties. In particular, mean path length was a dominant predictor of trophic levels, indicating that hierarchical position captures not only local asymmetries but also the efficiency with which each region integrates information within the effective connectivity network (Fig. 2d).

The directedness was closely associated with the asymmetry of causal interactions, as measured by the Frobenius norm in the GEC matrix. This highlights that the brain’s inherent out-of-equilibrium regime, driven by irreversible processes, promotes a more stratified, layered hierarchical configuration (Fig. 2e).

The DMN’s mediator position aligns with its known role as a high-level coordinator of internally guided cognition and cross-network integration (Raichle et al., 2001; Buckner et al., 2008; Smallwood et al., 2021; Deco et al., 2023). Its position at the centre of the hierarchy may also underlie its susceptibility to AD-related pathological spread, given its overlap with early tau burden.

### Stage-dependent hierarchical reorganization in AD

The hierarchical reorganization in the AD continuum was marked by both increased trophic levels in visual and thalamic regions and a decrease in motor, frontal, insula, hippocampal, and temporal regions. In the progression of the disease, these reductions extend to the cingulate, precuneus, and angular gyrus (Fig. 3a). At the network level, this progression is reflected in a transient shift in trophic levels, with an increase in VN and a decrease in SMN, SAN, and the DMN as pathology worsens. Interestingly, compared to HC,, trophic levels in MCI+ individuals showed a rebound to baseline before decreasing again in AD+, suggesting a reorganization before reaching later-stage hierarchical collapse (Fig. 3b).

This pattern maps directly onto the known propagation of AD pathology. Aβ accumulation begins in association cortices (Thal et al., 2002; Grothe et al., 2017), disrupting long-range integrative signalling, while tau spreads from the entorhinal cortex into hippocampal and DMN regions (Braak & Braak, 1991; Franzmeier et al., 2022; Schöll et al., 2016), weakening the functional leverage of these transmodal hubs. By contrast, early-preserved visual cortices maintain structural integrity longer into the disease course, consistent with their increased hierarchical position.

### Flattened hierarchy in AD

The directedness measure distinguished AD+ participants to have a more flattened hierarchy compared to all other groups, driven by lower trophic-level distances between regions (Fig. 3c). This indicates a decrease in the overall directionality of information flow within the brain’s causal network architecture, indicating more bidirectional, recurrent, and reversible dynamics. This pattern is consistent with the notion that AD brain dynamics are closer to equilibrium, characterized by less temporal asymmetry and reduced entropy production (Sun et al., 2020; Cruzat et al., 2023). Notably, this flattened hierarchical structure resembles the brain dynamics in states of reduced consciousness, such as sedation or deep sleep, where reduced levels of irreversibility lead to constrained computational capabilities (Luppi et al., 2024; Deco et al., 2022; García-Guzmán et al., 2023), in contrast to the higher non-equilibrium dynamics observed during cognitively demanding tasks associated with greater computational flexibility, allowing for more complex and adaptive information processing (Deco et al., 2023; Kringelbach et al., 2023). Thus, the hierarchical flattening in AD progression implies a compromised brain ability to process information and solve cognitive tasks, leading to an inefficient organization unable to support cognitive function.

This decreased verticality is partly compatible with interactions between Aβ and tau pathophysiology. Aβ-related hyperexcitability and hyperconnectivity (Busche et al., 2008; Palmqvist et al., 2017; Giorgio et al., 2024) facilitate tau propagation along connected networks (Jacobs et al., 2018; Adams et al., 2019; Roemer-Cassiano et al., 2025). The resulting destabilization of higher-order DMN and limbic circuits might lead to a reduction between trophic layers and contribute to an overall collapse in the brain’s hierarchical structure.

### Predicting AD stage using machine learning

Our machine-learning analysis further demonstrated that hierarchical features carry diagnostic information along the AD continuum. A logistic regression classifier reliably identified disease stages, including the subtle distinction between Aβ-negative and Aβ-positive healthy individuals (Fig. 3d). SHAP analyses revealed a coherent progression of hierarchical disruption, beginning with early subcortical alterations in HC+, extending to thalamic and fronto-striatal changes in MCI+, and culminating in widespread cortical–subcortical reorganization in AD+ (Supplementary Fig. 4).

### Associations with ATN biomarkers

Hierarchical alterations were systematically linked to ATN biomarkers (Fig. 4). Higher amyloid burden corresponded to increased trophic levels in the visual and subcortical regions and decreased levels in somatomotor, salience, and DMN networks. Tau pathology primarily affected the bilateral isthmus cingulate, left angular gyrus, and left hippocampus, while neurodegeneration (reduced GMV) strongly predicted lower trophic levels in hippocampus, DMN, and SAN, and higher levels in DAN. The DMN emerged as a common locus across all three biomarkers—a pattern fully consistent with its vulnerability to tau spread and its known coupling with amyloid pathology (Braak & Braak, 1991; Schöll et al., 2016; Busche & Hyman, 2020). Notably, the increase in trophic levels in VN and thalamus with amyloid burden further supports their selective vulnerability in later stages of AD s.

### Cognitive relevance of hierarchical disruption

Our hierarchical measures showed strong associations with cognitive decline across ADAS-Cog-13, CDR-SB, MoCA, and MMSE questionnaires. Poorer performance was linked to higher trophic levels in VN, and lower levels in SMN, SAN, CN, and DMN (Fig. 5a). Regionally, cognitive impairment corresponded to reduced trophic levels in somatomotor, frontal, angular gyrus, cingulate, and hippocampal regions, and increased levels in occipital and thalamus (Fig. 5b).

**Figure 5.**
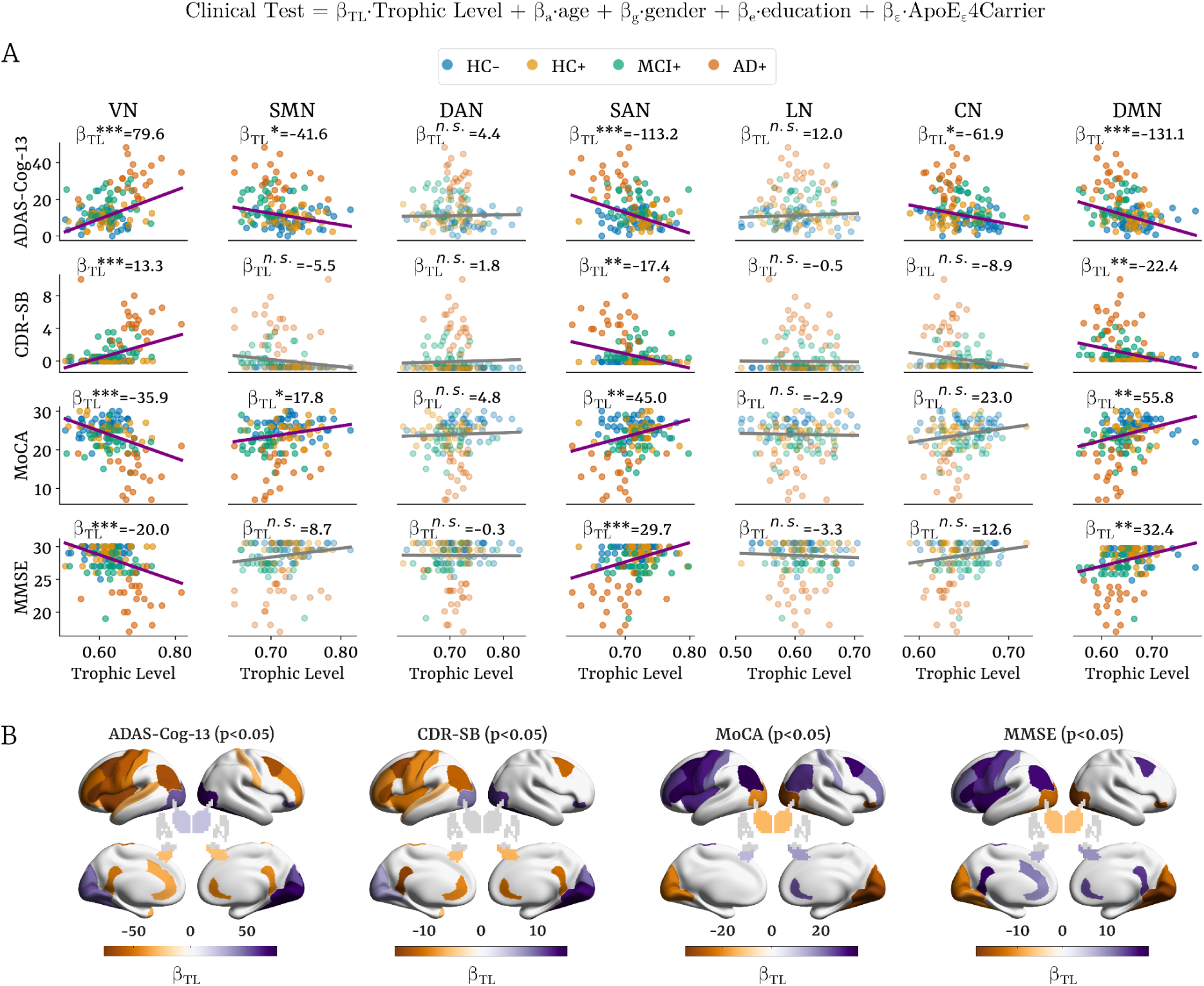
Trophic level associations with cognitive performance at network and regional level. **(A)** Regression plots illustrating the slope and significance of the associations between trophic levels in each network (columns) and clinical assessments (rows). **(B)** Regional brain renderings showing cortical and subcortical locations where trophic levels significantly predicted cognitive performance. Multiple linear regression adjusted for age, gender, education, and APOE-ε4 carrier status, and corrected for multiple comparisons using FDR. ADAS-Cog-13: Alzheimer’s Disease Assessment Scale – Cognitive-13, CDR-SB: Clinical Dementia Rating Sum of Boxes, MoCA: Montreal Cognitive Assessment, MMSE: Mini-Mental State Examination, VN: visual network, SMN: somatomotor network, DAN: dorsal attention network, SAN: salience network, LN: limbic network, CN: control network, DMN: default mode network.

These results highlight the relationship between hierarchical flattening and cognitive decline. The overall reduction in the brain’s hierarchical verticality reflects a less efficient and adaptive network, impairing the brain’s capacity to perform complex cognitive tasks, as evidenced by poorer clinical performance.

### Network reorganization in the AD continuum

The observed hierarchical alteration reflects a selective disruption of networks such as the DMN, SAN, and SMN, with compensatory shifts toward primary sensory-thalamic systems that remained relatively spared in the AD patients from this cohort. The DMN emerges as one of the earliest and most severely affected networks, with disruptions correlating with amyloid deposition and clinical severity (Palmqvist et al., 2017; Ozdemir et al., 2025). As pathology advances, both the DMN and SAN progressively decrease connectivity in preclinical stages of AD (Schultz. et al., 2017). Within this context of failure, reduced SAN influence–mediating between internally and externally directed cognition–and reduced SMN influence–suporting motor planning and execution–aligns with emerging evidence of impaired task-switching, attentional control and motor impairments in preclinical and mild AD preceding cognitive deficits (Buchman & Bennet, 2011; He et al., 2013; Zhang et al., 2020; Andrade-Guerrero et al., 2024).

In parallel, increased trophic levels in VN and thalamic regions indicate an increase in causal influence of less pathology-burdened circuits. This may reflect a maladaptive compensation, in which sensory and thalamic systems assume a greater role as cortical communication deteriorates. This partly aligns with evidence of VN hyperconnectivity related to visuospatial deficits in AD phenotypes and with findings that thalamic atrophy contributes to episodic memory impairment across clinical subtypes (Singh et al., 2023; Forno et al., 2023).

Overall, our findings support an interpretation of the trophic level rebalance in which: (1) early amyloid deposition in DMN hubs and subsequent tau-driven neurodegeneration lead to reduce causal influence of the DMN, SAN, and SMN; and (2) less protein-burdened regions such as visual and thalamic systems become more influential, likely using the spare capacity in these networks to maintain cortico-subcortical communication. This shift in the brain’s hierarchical organization provides a mechanistic bridge between ATN progression and the clinical profile in AD, where reductions in DMN, SAN and SMN influence and increases in visual and thalamic systems (Fig. 3a,b), correlates with amyloid burden (Fig. 4a) and cognitive decline (Fig. 5).

### Potential limitations

This study has several limitations. First, resting-state fMRI BOLD signals are indirect proxies of neural activity and incorporate vascular, metabolic, and physiological influences (Bandettini & Ungerleider, 2001; Logothetis, 2008). Although strongly correlated with local field potentials, they reflect mixed contributions from heterogeneous neuronal populations. Second, AD pathology is phenotypically diverse, with distinct tau-spread and atrophy subtypes (Murray et al., 2011; Poulakis et al., 2018). Larger samples will be necessary to capture subtype-specific hierarchical alterations. Finally, the cross-sectional design precludes causal inference about temporal progression.

### Conclusions

Using this novel ecology-inspired framework, we show that AD profoundly reshapes the hierarchical organization of brain dynamics. Across the disease continuum, AD is marked by elevated trophic levels in visual and thalamic systems and diminished levels in somatomotor, salience, control, and DMN regions, indicating a breakdown of the healthy, layered brain organization toward a more flattened hierarchy. This pattern reflects a shift towards a more reversible, less flexible, equilibrium-like dynamics, captured by reduced directedness, and tightly linked to ATN-dependent disruptions of causal interactions, resulting in cognitive decline across domains.

Our findings demonstrate that hierarchical organization—captured by trophic levels and directedness—provides a sensitive neuroimaging marker of the causal architecture underlying AD progression. This framework offers a mechanistic tool for disease staging, may inform therapeutic targeting aimed at restoring hierarchical flexibility and integration, and warrants future longitudinal validation to determine whether hierarchical changes precede clinical symptoms and can serve as early biomarkers.

## Methods

### Participants

The imaging and cognitive data involved in this study were obtained from the Alzheimer’s Disease Neuroimaging Initiative database (ADNI, available at https://adni.loni.usc.edu/). ADNI is a multi-center follow-up study from 2003, led by Principal Investigator Michael W. Weiner, dedicated to the development of multimodal biomarkers for the early detection and tracking of AD, that include clinical, imaging, genetic, and biochemical modalities.

Participants’ diagnostic status was determined according to ADNI’s established criteria (further details available at https://adni.loni.usc.edu/help-faqs/adni-documentation/). Four diagnostic groups were included: healthy controls devoid of amyloid pathology (HC−), with a Mini-Mental State Examination (MMSE) score ≥24, a Clinical Dementia Rating (CDR) of 0, and no signs of depression; healthy controls, amyloid-positive (HC+), meeting the same MMSE, CDR, and depression criteria; mild cognitive impairment, amyloid-positive (MCI+), defined by MMSE ≥24, CDR = 0.5, objective memory impairment based on the education-adjusted Wechsler Memory Scale II (D Wechsler, 1987), and preserved activities of daily living; and AD, amyloid-positive (AD+), characterized by MMSE scores between 20-26, CDR >0.5, and meeting the National Institute of Neurological and Communicative Disorders and Stroke/Alzheimer’s Disease and Related Disorders Association criteria for probable AD. Amyloid positivity was determined as amyloid centiloid-converted values >24. Details on how the Centiloid cutoff was computed and applied are provided in the *PET and Structural Data Processing* section of the *Methods*. More details in the MMSE are found in the Clinical Cognitive Assessments section.

We included 134 participants from the ADNI3 study, consisting of 46 HC−, 36 HC+, 31 with MCI+, and 21 with AD+. However, for analyses requiring clinical data along with baseline 18F-Florbetapir/Florbetaben amyloid-PET and 18F-Flortaucipir tau-PET scans, the number of subjects was reduced to 121: 41 with HC−, 34 with HC+, 27 with MCI+, and 19 with AD+. All subjects were right-handed. Details in age, gender, education, and other clinical characteristics are shown in Table 1. Originally, the MRI dataset consisted of 151 subjects, but during the preprocessing phase involved a two-step filtering process–scrubbing and harmonization (see Methods)–led to a reduction in the dataset size.

**Table 1.**
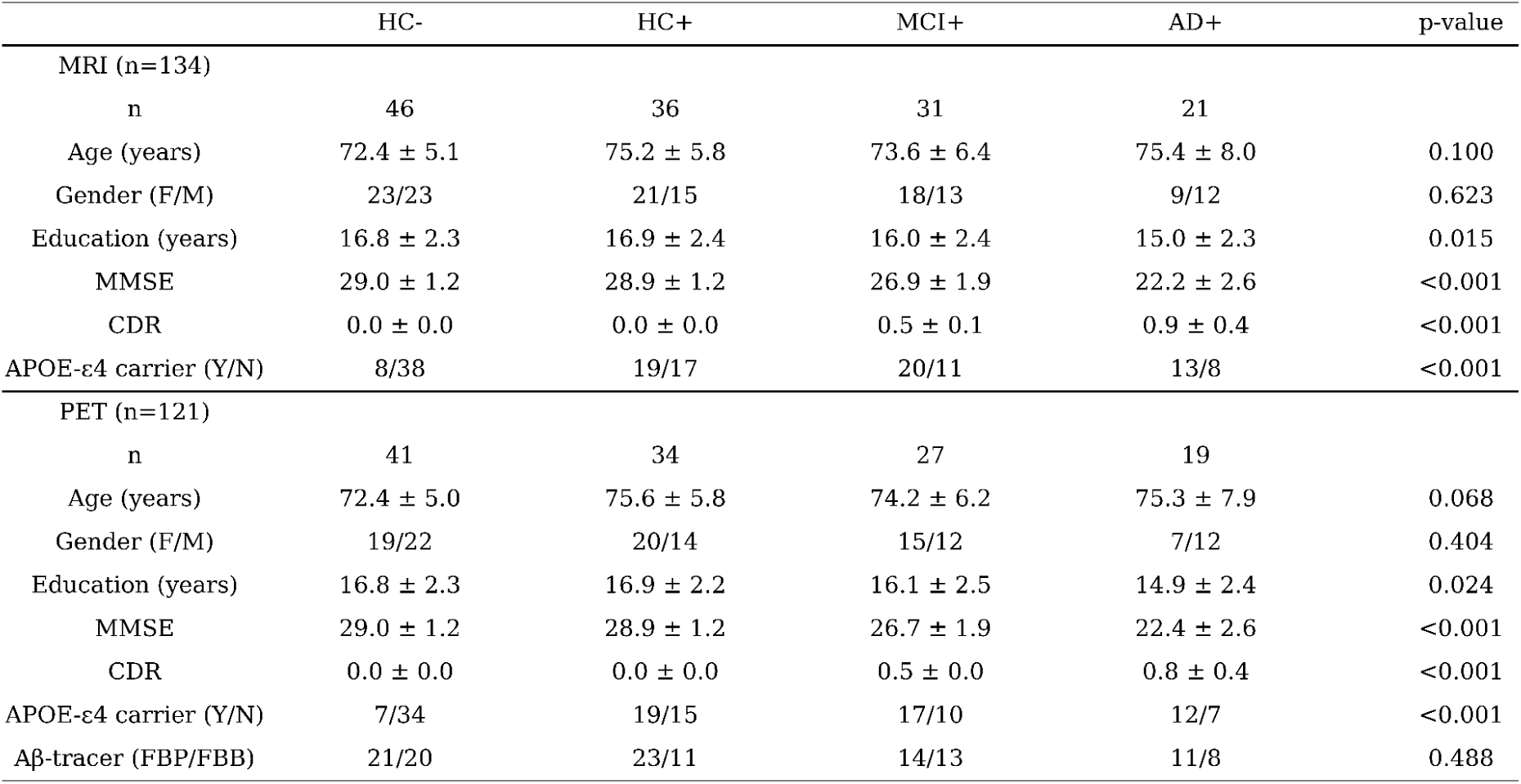
Demographics and clinical information. Data are presented as the means ± standard deviation (SD). *P-*values for age, education, MMSE, and CDR were calculated using the Kruskal-Wallis H test, while *p-*values for gender, APOE-ε4 carrier, and Amyloid tracer were obtained using a two-tailed Pearson chi-square test. FBP: 18F-Florbetapir; FBB: 18F-Florbetaben.

|  | HC- | HC+ | MCI+ | AD+ | p-value |
| --- | --- | --- | --- | --- | --- |
| MRI (n=134) |  |  |  |  |  |
| n | 46 | 36 | 31 | 21 |  |
| Age (years) | 72.4 ± 5.1 | 75.2 ± 5.8 | 73.6 ± 6.4 | 75.4 ± 8.0 | 0.100 |
| Gender (F/M) | 23/23 | 21/15 | 18/13 | 9/12 | 0.623 |
| Education (years) | 16.8 ± 2.3 | 16.9 ± 2.4 | 16.0 ± 2.4 | 15.0 ± 2.3 | 0.015 |
| MMSE | 29.0 ± 1.2 | 28.9 ± 1.2 | 26.9 ± 1.9 | 22.2 ± 2.6 | <0.001 |
| CDR | 0.0 ± 0.0 | 0.0 ± 0.0 | 0.5 ± 0.1 | 0.9 ± 0.4 | <0.001 |
| APOE-ε4 carrier (Y/N) | 8/38 | 19/17 | 20/11 | 13/8 | <0.001 |
| PET (n=121) |  |  |  |  |  |
| n | 41 | 34 | 27 | 19 |  |
| Age (years) | 72.4 ± 5.0 | 75.6 ± 5.8 | 74.2 ± 6.2 | 75.3 ± 7.9 | 0.068 |
| Gender (F/M) | 19/22 | 20/14 | 15/12 | 7/12 | 0.404 |
| Education (years) | 16.8 ± 2.3 | 16.9 ± 2.2 | 16.1 ± 2.5 | 14.9 ± 2.4 | 0.024 |
| MMSE | 29.0 ± 1.2 | 28.9 ± 1.2 | 26.7 ± 1.9 | 22.4 ± 2.6 | <0.001 |
| CDR | 0.0 ± 0.0 | 0.0 ± 0.0 | 0.5 ± 0.0 | 0.8 ± 0.4 | <0.001 |
| APOE-ε4 carrier (Y/N) | 7/34 | 19/15 | 17/10 | 12/7 | <0.001 |
| Aβ-tracer (FBP/FBB) | 21/20 | 23/11 | 14/13 | 11/8 | 0.488 |

A detailed characterization of ATN biomarkers–including Aβ, tau, and grey matter volume (GMV) for each stage–is provided in the Supplementary Material (see *ATN Biomarkers for Neuropathological Change, Supplementary Figure 1*), describing their cortical and subcortical spatial burden accumulation, patterns of atrophy progression, and network-level changes across the AD continuum.

All study procedures were conducted in accordance with the Declaration of Helsinki, and ethical approval was obtained by the ADNI investigators. All the participants provided written informed consent.

### Neuroimaging Acquisition

Structural, resting-state functional MRI (rs-fMRI), and diffusion-weighted (DWI) images were obtained from all sites using a 3-Tesla Siemens and GE scanners. T1-weighted structural scans were collected using an MPRAGE sequence (TR/TE=2300/3ms; Voxel size=1×1×1mm). rs-fMRI was obtained using a 3D echo-planar imaging (EPI) sequence with 197 fMRI volumes per subject (TR/TE=3000/30ms; flip angle=90°; Voxel size=3.4×3.4×3.4mm). In multi-shell multiband dMRI (TR=3400ms; Voxel size=2×2×2mm), protocols varied in the number of DWI directions (i.e., angular resolution), and the number of non-diffusion sensitized gradients (*b_0_* images). PET data were assessed post intravenous injection of 18F-labeled tracers (Flortaucipir: ×x5min time-frames, 75-105min post-injection; Florbetapir: 4×5min time-frames, 50-70min post-injection; Florbetaben: 4×5min time-frames, 90-110min post-injection). Detailed imaging protocols and scanning parameters can be found in ADNI (https://adni.loni.usc.edu/help-faqs/adni-documentation/).

### Clinical Cognitive Assessments

We selected cognitive assessments based on their clinical relevance and widespread use in dementia research, prioritizing those available for the largest number of participants in our dataset. These included the previously mentioned MMSE (Folstein et al., 1975) and CDR (Berg, 1989), as well as the Alzheimer’s Disease Assessment Scale – Cognitive-13-item version (ADAS-Cog-13; Rosen et al., 1984), and the Montreal Cognitive Assessment (MoCA; Nasreddine et al., 2005). This selection of cognitive assessments was intended to capture a broad range of cognitive domains, including global cognition, memory, language, executive functioning, and daily functional abilities.

For each participant, we included the cognitive assessments corresponding to the same study phase as the MRI data (ADNI3) and available for the same individuals in the demographics dataset. Only baseline or screening visits (e.g., *bl*, *sc*) were kept, ensuring that the cognitive tests were conducted closest to the corresponding imaging session. In cases where there were multiple tests for the same participant, the earliest visit was selected. Additional details on the cognitive assessments can be found in the Supplementary Material (see *Clinical Assessments Information*), and protocols in the ADNI documentation (https://adni.loni.usc.edu/help-faqs/adni-documentation/).

### PET and Structural Data Processing to Obtain ATN Biomarkers

PET images were downloaded from the ADNI database after quality control by the PET Core Group (Jagust et al., 2024). Preprocessing was performed in the Statistical Parametric Mapping (SPM, version SPM25), which included co-registration of each PET image to the native T1-weighted MRI, which was then segmented to obtain the deformation fields used for the normalization of the PET scans to MNI-152 standard space with 2mm isotropic resampling resolution. PET scans were smoothed with a Gaussian kernel whose FWHM was chosen for each scan so that the resulting effective image resolution was 8mm FWHM. Partial volume correction was applied after the co-registration step using the Müller-Gärtner algorithm implemented in the PETPVC toolbox (Thomas et al., 2016). Following the standard ADNI PET pipeline, all PET images were converted to Standardized Uptake Value Ratios (SUVRs) using the whole cerebellum as a reference for amyloid tracers, Florbetapir and Florbetaben (Bullich et al., 2017; Cho et al., 2020), and the inferior cerebellar gray matter for the tau tracer, Flortaucipir (Baker et al., 2017; Young et al., 2021). Both Florbetapir and Florbetaben SUVRs were converted to Centiloid (CL) units using specific calibration equations (Florbetaben: CL = 94.082 × SUVR – 94.772; Florbetapir: CL = 110.982 × SUVR – 116.172), following the GAAIN Centiloid Project Level-1 and Level-2 procedures (Klunk et al., 2015).

The single CL value for each participant to determine Aβ positivity was computed by averaging the SUVR within the global cortical target (CTX) region’s set (Klunk et al., 2015), and then converting this value to CL units. The CTX volume-of-interest (VOI), as defined by Klunk et al. (2015), was data-driven by capturing cortical areas with the greatest amyloid load in typical AD subjects and age-matched controls. This VOI includes frontal, temporal, and parietal cortices, precuneus, anterior striatum, and insular cortex. Subjects with a single CL value in the CTX above 24 were classified as Aβ-positive; otherwise, they were considered Aβ-negative. This threshold was established in Navitsky et al. (2018) as the cutoff that best discriminated cases with moderate to frequent neuritic plaques from those with none or sparse plaques in autopsy-confirmed data.

Structural T1-weighted images were preprocessed using fMRIPrep (version 24.1; Esteban et al., 2019), which incorporates FreeSurfer’s (v7.4.1) recon-all pipeline (Dale et al., 1999; Fischl et al., 1999) for cortical surface reconstruction and volumetric segmentation that includes skull stripping, volumetric labelling, bias correction, tissue segmentation into grey matter, white matter, and cerebrospinal fluid, cortical surface extraction, and parcellation. GMV is represented as the sum of voxels within each anatomical region of interest, in mm³.

### APOE genotyping

Genetic samples were collected in 10 mL ethylenediaminetetraacetic acid (EDTA) blood tubes at the baseline visit and then analyzed at the National Centralized Repository for Alzheimer’s Disease (NCRAD) to identify ε2, ε3, and ε4 alleles. Participants carrying one or two ε4 alleles were classified as ε4-carriers; all others were classified as non-carriers.

### fMRI and DWI Data Preprocessing

The fMRI dataset was organised in the Brain Imaging Data Structure (BIDS) standard so as to be passed to fMRIPrep (version 24.1; Esteban et al., 2019). Detailed steps and functions are provided in the Supplementary Material (see *Image Pre-processing with fMRIPrep*). The DWI dataset was preprocessed using MRtrix3 and FSL tools (Tournier et al., 2019; Smith et al., 2004). Detailed preprocessing steps and commands are provided in the Supplementary Material (see *Derivation of Structural Connectomes from Diffusion MRI*).

### Regressing Confounds, Brain Parcellation, and Network-Level Computation

To reduce potential confounding effects in the fMRI data, several regressors were included in analyses. First, we applied a high-pass filter to remove low-frequency signals that can be introduced by physiological and scanner noise sources. Next, to reduce the effects of head motion, we regressed out the six rigid-body motion parameters, which have been previously demonstrated to introduce bias in group comparisons (Power et al., 2012). These motion parameters include the transition on the three axes (x, y, z) and the respective rotation (α, β, γ), which are estimated relative to a reference image (Friston et al., 1996).

Additionally, to minimize the impact of non-neuronal BOLD signal fluctuations, which are unlikely to reflect anatomical activity (Fox et al., 2005), we regressed out signals from white matter and cerebrospinal fluid. These signals, as well as the motion parameters, were expanded using the first temporal derivatives and their quadratic terms to capture potential non-linear effects of these noise sources (Satterthwaite et al., 2013). Furthermore, we removed high-motion segments in which the framewise displacement (FD) exceeds 0.5 mm, a process known as scrubbing (Power et al., 2012). FD measures the movement of the head from one volume to the next, helping to identify and exclude volumes with excessive motion. Finally, we included the standardized DVARS, defined as the root mean squared intensity difference between two consecutive volumes, and applied a threshold of 3 to identify and exclude volumes with excessive signal intensity changes. These regression steps were based on a denoising strategy proposed by Wang *et al*. (2024), which aims to improve the quality of fMRI connectivity studies. During this process, subjects with fewer than 150 time points were excluded due to their significantly lower signal length, in contrast to the subjects without scrubbing applied, which had 197 time points.

This regression procedure was applied to the corresponding preprocessed NifTI images through a brain atlas for denoising. Although there is no consensus on the optimal spatial brain parcellation (Eickhoff et al., 2018), all neuroimaging data in this study were preprocessed using the DK80 parcellation. The DK80 combines the Mindboggle-modified Desikan Killiany parcellation (Desikan et al., 2006) with a total of 62 cortical regions (32 regions per hemisphere) (Klein & Tourville, 2012) and 18 subcortical regions (9 per hemisphere): hippocampus, amygdala, thalamus, putamen, caudate, nucleus accumbens, subthalamic nucleus, globus pallidus internal segment, and globus pallidus external segment. This parcellation was chosen based on previous work that showed its effectiveness for whole-brain modeling when measuring brain hierarchy (Deco et al., 2024; Deco et al., 2021). To provide a clearer and concise visualization of the subcortical results, we presented a single 2D slice in MNI152 space (y=-8mm), omitting an additional slice that would include the nucleus accumbens, as this region did not reveal relevant findings in the present study.

In addition to the region-level parcellation, we also considered a network-level parcellation dividing the brain into seven functional networks based on the Schaefer atlas (Schaefer et al., 2018): visual network (VN), somatomotor network (SMN), dorsal attention network (DAN), salience network (SAN), limbic network (LN), control network (CN), and default mode network (DMN). Although Ji et al. (2019) assigned subcortical voxels to cortical networks based on the strongest functional connectivity, the subcortical regions appear to show connections with multiple cortical networks—for instance, the thalamus couples with VN, SMN, DAN, CN, and DMN (Chumin et al., 2022)—and including those regions would therefore introduce overlap and affect the interpretation of the metrics. To determine which cortical regions corresponded to each functional network, we overlapped the regional-level DK80 atlas with the network-level Schaefer atlas, assigning each region to the network with which it shared the greatest voxelwise overlap. The purpose of including these functional networks, further from reducing the dimensionality and statistical comparisons, was to provide a more direct analysis of cognitive domains rather than pure anatomical parcellation, allowing for a more concise analysis of brain changes in AD (Figure 1C).

In this study, network-level values were obtained by averaging the regional measures (Aβ, tau, GMV, and trophic levels) across all regions belonging to each functional network. The mean, rather than the sum, was used to compute network-level values to ensure that each region contributed equally to the corresponding network. Using the mean provides a measure independent of size, whereas summing regional values would weight larger regions or networks with more parcels in a disproportional way. This approach was applied across all analyses in this study.

### Harmonization

When collecting MRI data from multiple sites, bias and variability–unrelated to biology–arise. To correct this, harmonization procedures are essential to make data more comparable across scanners by making sure that important biological variables (age, gender, and diagnosis) remain preserved when removing scanner effects. In this study, we used ComBat, a popular and reliable harmonization tool that has demonstrated high accuracy in eliminating site effects (Yu et al., 2018; Bell et al., 2022) compared to other methods, such as the traveling-subject approach (Roffet et al., 2022). ComBat was originally proposed by Johnson et al. (2007) as a batch-effect correlation tool for genomics data, which was later adapted for neuroimaging by Fortin et al. (2017) to regress out effects induced by site in MRI data. ComBat operates within an empirical Bayes framework to fit a set of covariates to the data and then applies a linear shift-and-scale transformation to the model residuals to align them with a reference distribution. The covariate effects are then reintroduced, yielding an adjusted set of data for each site while retaining the influence of the biological variables. ComBat uses a multivariate linear mixed effects regression with terms for biological variables and scanner to model imaging feature measurements as follows:

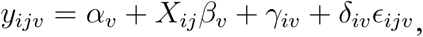

where *y_ijv_* represents the brain measure (e.g., trophic level, amyloid/tau accumulation, GMV, etc.) at site *i* and participant *j* of the feature *v* (i.e., region or network), *α_v_* is the average across participants of the measure, *X_ij_* is the design matrix for the covariances of interest (age, gender, education and diagnosis), and *β_v_* is a vector of regression coefficients corresponding to *X*. The terms *γ_iv_* and *δ_iv_* represent the additive and multiplicative site effects of site *i* for the feature *v*, respectively, while*∊_ijv_* represents an error term assumed to arise from a normal distribution with zero mean and variance 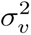. The ComBat-Harmonized brain measures are then defined as:

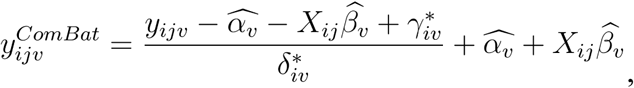

Where 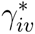 and 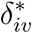 are the empirical Bayes estimates of *γ_iv_* and *δ_iv_*, respectively.

Data harmonization was implemented in Python using the *harmonizationLearn* and *harmonizationApply* functions from the *neuroHarmonize* package (Pomponio et al., 2020), using its default linear ComBat implementation (i.e., without the non-linear ComBat-GAM extension), with covariates of age, gender, education, and diagnosis. Subjects with unique sites were excluded from the harmonization process, as harmonization methods rely on data with multiple sites for effective calibration. To test that the quality of the harmonization was accurate for our dataset, we performed two independent analyses: (1) verifying that the expected age-related neurodegeneration in GMV (i.e., lower values with increasing age) persisted after harmonization, and (2) confirming that significant trophic levels differences between sites within group were eliminated. Both tests succeeded (see Supplementary Figures 5 and 6). Harmonization was then computed separately for trophic levels, directedness, Aβ, tau, and GMV at the regional level, before averaging within networks.

### GEC Model Optimization - The Hopf Model

To compute the trophic levels, a directed graph representing causal interactions between brain regions is estimated by whole-brain modeling of each individual’s resting state, combining anatomical connectivity with local dynamics simulated using the Hiopf model in order to fit the dynamics of empirical neuroimaging data (Deco & Kringelbach, 2014; Kringelbach & Deco, 2020; Breakspear, 2017; Figure 1A).

The Hopf model emulates the dynamics emerging from the mutual interactions between brain areas, considered to be interconnected on the basis of anatomical structural connectivity (Deco et al., 2017a). In other words, it combines brain structure and activity dynamics to explore and explain the underlying mechanisms of brain connectivity. In this study, we employed the Stuart-Landau oscillator model (first introduced by Matthews & Strogatz, 1990) to represent the local dynamics of each brain area described by the normal form of a supercritical Hopf Bifurcation (Supplementary Figure 7). This is the canonical model for studying the transition from noisy, asynchronous, damped oscillations to oscillatory, synchronous, self-sustained oscillations (Kuznetsov, 1998). The interaction between different Hopf oscillators using brain network architecture has been shown to successfully replicate features of brain dynamics observed in EEG (Freyer et al., 2011, 2012), MEG (Deco et al., 2017b) and fMRI (Deco et al., 2017a; Deco et al., 2019; Deco et al., 2022; Kringelbach et al., 2023; García-Guzmán et al., 2023; Deco et al., 2023) and, more recently, to determine hierarchical configurations in the brain (Deco et al., 2024). The full mathematical description of the Hopf model and its linearization is available in the Supplementary Material (see *The Hopf Model* subsection from *Generative Effective Connectivity Methodology*).

To model the whole-brain dynamics requires modelling the coupling of the local dynamics of each brain region interconnected through a given coupling connectivity matrix, *C*. Here, we use a pseudo-gradient procedure to optimize *C*. Initially, the structural connectivity matrix corresponded to the individual’s group-average connectome, computed from the diffusion MRI data; the full processing pipeline is detailed in the Supplementary Material (*Derivation of Structural Connectomes from Diffusion MRI*). The final optimized matrix comprises the effective connectivity values for each anatomical pair connection; i.e., we iteratively compared the output of the model with empirical measures of the FC matrix (*FC^empirical^*), that is, the normalized covariance matrix of the functional neuroimaging data.

Additionally, the break of symmetry to capture causal dynamics was then computed by fitting the non-reversibility. We define the forward and reversed matrices of time-shifted correlation for the forward version and the respective reversed backward version of each brain region’s time series, i.e., reversing in time the natural forward evolution of the time series. This allows us to compute both forward and reverse matrices, expressing the functional causal dependencies between regions for the forward and artificially generated backward version. We compared the output of the model with the forward normalized *τ* time-shifted covariances 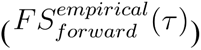. The normalized time-shifted covariances were derived by shifting the empirical covariance matrix 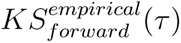 and dividing each pair (*i*, *j*) by 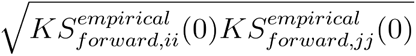. To fully capture the asymmetry, we fit the non-reversibility by performing the same procedure on the reversed normalized *τ* time-shifted covariances 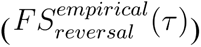.

The updated heuristic pseudo-gradient algorithm follows the form:

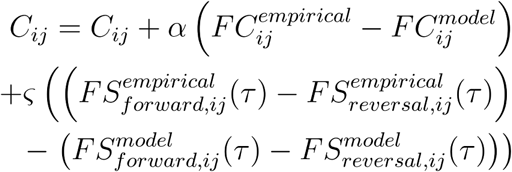

Where 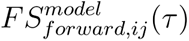 is defined similarly as for forward 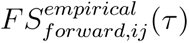, and 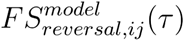 as for reversal 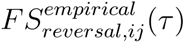. In other words, for the forward version, it is given by the first *N* rows and columns of the simulated *τ* time-shifted covariances 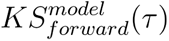 normalized by dividing each pair (*i*, *j*) by 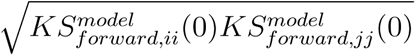, where the 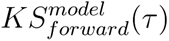 is the shifted simulated covariance matrix computed as

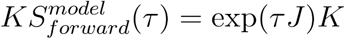

where the *J* matrix is the Jacobian at the fixed point, where the linearization of the Hopf model is valid. The same procedure was applied to the reversal version of 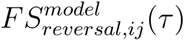. The model is executed iteratively with the updated *C* until a stable and convergent fit is achieved. We use this method in two stages: first, at the group level, which then, second, is used as the starting point for individual optimization. In summary, we refer to the optimized matrix *C* as the GEC. The full mathematical description of the optimization procedure is available in the Supplementary Material (see *Linearization of the Hopf Model* subsection from *Generative Effective Connectivity Methodology*).

This framework–an extension of the classic effective connectivity (Friston et al., 2003) but generative in the sense that the strengths of existing anatomical connectivity are fine-tuned to fit empirical data–is computationally more efficient than those including hemodynamics, such as the Balloon model (Friston et al., 2000) in dynamic causal modelling (DCM) (Friston et al., 2003), Granger causality (Granger, 1980) and transfer entropy (Schreiber, 2000). Directionality of information flow was measured using a thermodynamic-inspired approach by quantifying the level of asymmetry between forward and backward interactions, i.e., non-reversibility, capturing the inside-out balance of intrinsic and extrinsic brain dynamics (Deco et al., 2022; Deco et al., 2023). This asymmetry in time that leads to the breaking of the detailed balance reflects the arrow of time, which, in turn, quantifies the extent to which one region is driving another, pushing away the system from equilibrium toward a non-equilibrium state, resulting in a hierarchical organization (Kringelbach et al., 2023). This improves functional connectivity by approximating the asymmetrical causal impact between brain regions and thus results in a directed and weighted network representation (Deco & Kringelbach, 2014; Kringelbach & Deco, 2020). This framework reduces the degrees of freedom for each brain region to simplify non-linear dynamics while structural data excluded non-existing connections, thereby facilitating the analysis of larger networks and whole-brain parcellations in contrast to the limited number of regional ranges given by DCM, although recent work has applied DCM to the whole brain (Frassle et al., 2017; Prando et al., 2020; Razi et al., 2017).

### Functional Hierarchy Metrics - Trophic Levels and Directedness

The GEC defines a directed weighted graph of *N* nodes determined by the elements of *C*, where *w_mn_* describes the influence (weight) of *m* over *n*. For each node *n* the in-weights 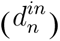 and the out-weights 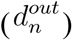 are given by the sum of the *m* columns and rows of *C,* respectively:

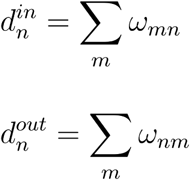

The total weight of node *n* is then defined by

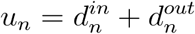

and the imbalance by the difference between the flow out and into the node n

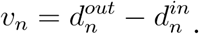

The graph-Laplacian Δ will be given in matrix form

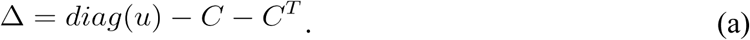

Hence, the trophic level is the solution vector *h* of the linear system of equations (Carmel et al., 2002)

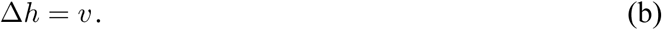

From this equation, we can infer that the trophic levels *h* are determined by the imbalance *v*. Indeed, trophic levels organize each region along the global directed flow of influence, revealing the functional hierarchical organization of the network–from sources of causal influence (*v* > 0), to mediators balancing inflow and outflow (*v* ≈ 0), to sinks where information converges (*v* < 0). Importantly, trophic levels are not exclusively derived from the local measures of a node’s balance but reflect the entire pattern of directed influence across the network. To further interpret this measure, we investigated its relation with other network metrics (see next section), revealing a significant correlation with communication efficiency (see Results section).

Once the first node’s trophic level is computed, this will be treated as the baseline for the hierarchical structure of the graph. Then, the trophic level of a given node indicates its position in the hierarchy relative to this baselone node. Positive values indicate nodes that are above in the hierarchy, and negative values indicate nodes that are below in the hierarchy. Here, we will be shifting all values by the smallest trophic level so that we get positive values for the overall hierarchical distribution of our graph. Consequently, the bottom of the hierarchy will be given by the node with a zero trophic level.

We complement the characterization of the network’s hierarchical organization with a global measure of directedness (MacKay et al., 2020). In a perfect coherent network with an ideal hierarchy, every edge increases the trophic level by 1, *h_i_* – *h_j_* = 1, and the directedness is maximal. To quantify the deviations from this ideal configuration (*∊_ij_ = h_i_* – *h_j_* – 1), we compute the overall incoherence of the network by

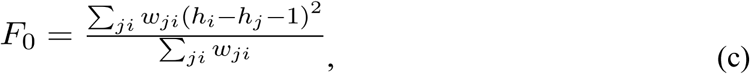

where the weight *w_ji_* ensures that stronger edges contribute more to the measure, which is normalized for independence of the scale of the graph. We then define the trophic directedness to be 1 - *F*_0_ so that the network will be considered as incoherent if *F*_0_ = 1 and fully coherent if *F*_0_ = 0. Hence, a flat hierarchy will be characterized by equal trophic levels and low directedness, reflecting a low level of directionality of information flow (high symmetry; Deco et al., 2022). In contrast, a strong vertical hierarchy will be associated with high directedness and strong asymmetric connections in a multiple-layered network.

It is worth noting that equations (a) and (b) can be derived directly by first writing equation (c) for generic levels, and then finding the solution *h* that minimizes *F*, which leads to Equation (b). In other words, the trophic levels are those that minimize the trophic incoherence (maximizing directedness). A complete mathematical explanation on how the trophic levels are calculated by minimizing a hierarchy energy function, resulting in equation (b), is available in the Supplementary Material (see *Hierarchy Metrics: Trophic Levels*).

Directionality and hierarchical orchestration could be captured in many ways, by reciprocity (proportion of bidirectional connections), or non-normality (overall asymmetry of the network). However, both reciprocity and non-normality lack sensitivity to the flow of connectivity. Instead, trophic directedness is a more structurally informative measure assigning trophic levels to each node of the network to reveal the tendency of edges to align in a global direction, i.e., the travel of information in one direction (see Supplementary Figure 8).

### Machine Learning Pipeline

We applied a machine learning (ML) framework to assess whether disease stage (HC−, HC+, MCI+, AD+) could be classified using the regional and network-level trophic levels and directness as input features. Independent binary classification problems were implemented to dissociate the diagnostic groups under study (HC− vs. HC+, HC− vs. MCI+, MCI+ vs. AD+, HC− vs. AD+). All analyses were conducted using the scikit-learn Python library (Pedregosa et al., 2011). Logistic Regression was used as the base classifier (Hosmer et al., 2013). To reduce dependence on a single data split and obtain more robust performance estimates, we performed N = 20 Monte Carlo iterations of the full model training and evaluation pipeline (Xu & Liang, 2001).

For each iteration, the dataset was divided into a 75% training split and a 25% test split, stratified by diagnostic group to preserve class proportions (Pedregosa et al., 2011). The training portion was used to perform a three-dimensional grid search for hyperparameter optimization using nested Leave-One-Out Cross-Validation (LOOCV), an approach that provides nearly unbiased generalization estimates for small datasets (Varma & Simon, 2006). The explored hyperparameters included: (i) number of selected features (NF), (ii) inverse regularization strength (C), and (iii) penalty type (L1 or L2; Table 2). For each hyperparameter combination, LOOCV was performed by iteratively training the model on M−1 samples and validating on the held-out sample, where M denotes the number of subjects in the training split. Performance across all folds was aggregated, and the hyperparameter set yielding the highest Area Under the ROC Curve (ROC–AUC) was selected as optimal.

**Table 2.** Hyperparameters explored for model tuning.

| Hyperparameter | Explored Values |
| --- | --- |
| NF | 5-15 |
| C | 0.01, 0.1, 1, 5, 10, 50, 100, 500, 1000 |
| Penalty Function | L1, L2 |

**Table 3.**
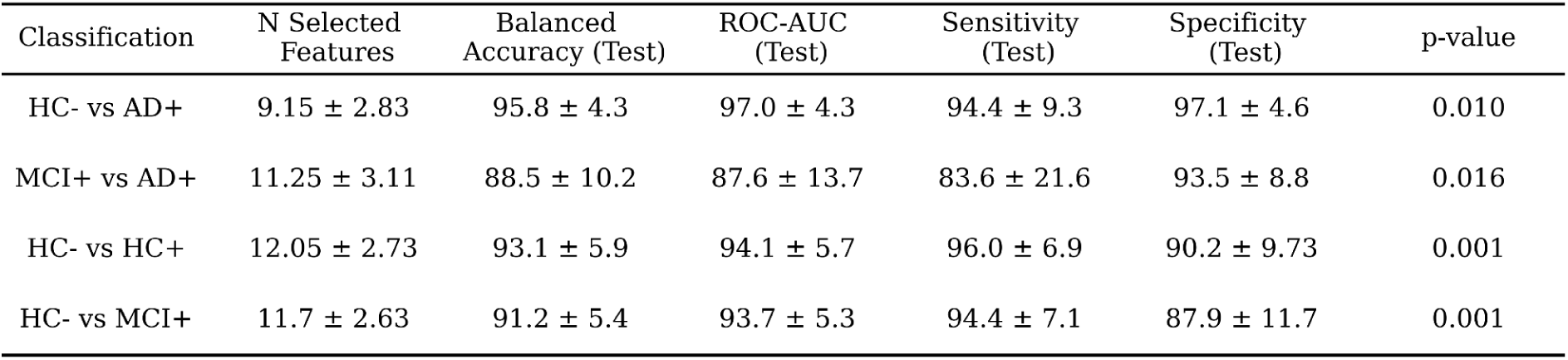
Best classification metrics for trophic levels features across comparisons.

| Classification | N Selected Features | Balanced Accuracy (Test) | ROC-AUC (Test) | Sensitivity (Test) | Specificity (Test) | p-value |
| --- | --- | --- | --- | --- | --- | --- |
| HC- vs AD+ | $9.15 \pm 2.83$ | $95.8 \pm 4.3$ | $97.0 \pm 4.3$ | $94.4 \pm 9.3$ | $97.1 \pm 4.6$ | 0.010 |
| MCI+ vs AD+ | $11.25 \pm 3.11$ | $88.5 \pm 10.2$ | $87.6 \pm 13.7$ | $83.6 \pm 21.6$ | $93.5 \pm 8.8$ | 0.016 |
| HC- vs HC+ | $12.05 \pm 2.73$ | $93.1 \pm 5.9$ | $94.1 \pm 5.7$ | $96.0 \pm 6.9$ | $90.2 \pm 9.73$ | 0.001 |
| HC- vs MCI+ | $11.7 \pm 2.63$ | $91.2 \pm 5.4$ | $93.7 \pm 5.3$ | $94.4 \pm 7.1$ | $87.9 \pm 11.7$ | 0.001 |

Feature selection was performed exclusively on the training data to avoid information leakage. For each candidate value of NF in the grid search, we applied the mRMR algorithm to identify the top NF non-redundant and maximally informative features (Peng et al., 2005). This procedure reduces dimensionality, improves computational efficiency, and mitigates redundancy among predictors.

Following hyperparameter optimization, a Logistic Regression model with the selected parameters was trained on the full training split. ROC curves were generated by computing the True Positive Rate (TPR) and False Positive Rate (FPR) across probability thresholds. The final decision threshold was chosen to maximize Youden’s J statistic (J = sensitivity + specificity − 1; Youden, 1950), where a sample was classified as positive if its predicted probability satisfied *P*(*x*) ≥ *threshold*. The held-out test split was then used to generate predictions and evaluate classification performance using balanced accuracy, ROC–AUC, sensitivity, specificity, and F1-score. Statistical significance of test accuracy was assessed using permutation testing, a non-parametric method for inference in ML classification (Itälinna et al. 2023). For each iteration, we compared the observed accuracy to a null distribution derived from 1,000 random permutations of the test labels; significance was defined as p < 0.05.

Performance metrics were aggregated across the N Monte Carlo iterations. For accuracy, ROC–AUC, sensitivity, and specificity, we report the mean and standard deviation across the N test splits. For permutation testing, the median p-value and interquartile range (IQR) across iterations are provided. The average ROC curve was computed using vertical averaging (Fawcett, 2006). Confusion matrices were averaged across iterations, with cell values representing the proportion of true labels predicted as each class.

Model explainability was assessed using SHAP (SHapley Additive exPlanations), a unified framework for interpreting model predictions based on Shapley values (Aas et al. 2021). Feature importance was quantified as the mean absolute SHAP value per feature. Because NF varies across iterations and selected feature subsets differ, features not chosen in a given model were assigned an importance value of zero. SHAP values were then averaged across iterations to obtain a robust measure of feature relevance.

### Trophic Level’s Relationship with Graph Metrics

In the previous section, we described how trophic levels are computed as a measure of hierarchical organization based on the directionality in the network’s topology. To further characterize the trophic levels as a network measure, we examined its associations with a range of standard graph metrics describing different topological properties computed from each individual’s GEC. Initially, we considered a set of structural and centrality measures, including in-degree, out-degree, out-in-degree ratio, closeness centrality, PageRank, clustering coefficient, mean path length, and betweenness centrality. To avoid redundancy due to collinearity, we excluded measures to avoid overlapping. The final analysis focused on out-in-degree centrality ratio, clustering coefficient, mean path length, and betweenness centrality, which together capture distinct aspects of the network’s topological characteristics. The equations used in this analysis are available in the Supplementary Material (see *Graph Metrics*).

We conducted regression analyses to identify which of these metrics significantly predict trophic levels by implementing ordinary least squares regressions, where each metric represents an independent variable and the trophic level is the dependent variable. Before fitting, all measures were z-scored. The resulting coefficient and significance revealed the contributions of each network property to the trophic level.

To assess the relationship between directedness and asymmetry in the causal interactions given by the GEC matrices, we computed asymmetry using the Frobenius norm. For each GEC matrix *A*, we determined the asymmetry index as the ratio of the Frobenius norm of the antisymmetric 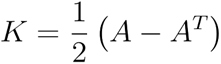 component of *A*, to the Frobenius norm of the full matrix *A*, 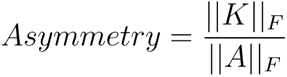. This measure captures the relative contribution of asymmetric interactions. Values close to 0 indicate predominantly symmetric interactions, whereas values close to 1 indicate highly asymmetric.

### Statistical Analysis

Group comparisons (HC−, HC+, MCI+, and AD+) were assessed at both regional and network levels using non-parametric permutation tests with covariate adjustment for age, gender, and education. To account for multiple comparisons, a False Discovery Rate (FDR) correction was applied. The variance in trophic levels explained by Aβ, tau, and GMV across brain regions and networks was evaluated using mixed-effects with ridge regression–a regression method that stabilizes coefficient estimation in the presence of multicollinearity between predictors, such as that occurring in this study between biomarkers. Here, age, gender, education, and APOE-ε4 carrier status were included as covariates, and a random intercept for subject was modeled to account for within-subject variability; permutation testing was computed for the validation of statistical significance. Previous to model fitting, continuous predictors (Aβ, tau, GMV, age, and education) were standardized due to their different numerical ranges to place them on a comparable scale, necessary for ridge regression. The predictor-outcome relationship was inverted to assess whether hierarchy predicted clinical test scores using multiple linear regression while adjusting for age, gender, education, and APOE-ε4 carrier status, and correcting for multiple comparisons using FDR.

It may be argued that the group should be included as a covariate in both regression models. However, in the first case (ridge regression), the disease stage is strongly driven by Aβ deposition, tau burden, and atrophy—as discussed extensively in the literature and in the introduction—so including this predictor would likely reduce the specific contributions of these biomarkers to trophic levels. In the second case, where trophic level measures are used to predict cognitive performance, the group could not be included as an independent variable because it is inherently defined by the cognitive assessments themselves.

## Data Availability

All data produced in the present study are available upon reasonable request to the authors.
Data used in preparation of this article were obtained from the Alzheimer′s Disease Neuroimaging Initiative (ADNI) database (https://adni.loni.usc.edu/).

https://adni.loni.usc.edu/

## Acknowledgements

Data collection and sharing for this project were funded by the Alzheimer’s Disease Neuroimaging Initiative (ADNI) (National Institutes of Health Grant U01 AG024904) and DOD ADNI (Department of Defense award number W81XWH-12-2-0012). ADNI is funded by the National Institute on Aging, the National Institute of Biomedical Imaging and Bioengineering, and through generous contributions from the following: Alzheimer’s Association; Alzheimer’s Drug Discovery Foundation; BioClinica, Inc.; Biogen Idec Inc.; Bristol-Myers Squibb Company; Eisai Inc.; Elan Pharmaceuticals, Inc.; Eli Lilly and Company; F. Hoffmann-La Roche Ltd and its affiliated company Genentech, Inc.; GE Healthcare; Innogenetics, N.V.; IXICO Ltd.; Janssen Alzheimer Immuno therapy Research & Development, LLC.; Johnson & Johnson Pharmaceutical Research & Development LLC.; Medpace, Inc.; Merck & Co., Inc.; Meso Scale Diagnostics, LLC.; NeuroRx Research; Novartis Pharmaceuticals Corporation; Pfizer Inc.; Piramal Imaging; Servier; Synarc Inc.; and Takeda Pharmaceutical Company. The Canadian Institutes of Health Research is providing funds to support ADNI clinical sites in Canada. Private sector contributions are facilitated by the Foundation for the National Institutes of Health (https://fnih.org/). The grantee organization is the Northern California Institute for Research and Education, and the study is coordinated by the Alzheimer’s Disease Cooperative Study at the University of California, San Diego. ADNI data are disseminated by the Laboratory for Neuro Imaging at the University of California, Los Angeles.

Data used in preparation of this article were obtained from the Alzheimer’s Disease Neuroimaging Initiative (ADNI) database (https://adni.loni.usc.edu/). As such, the investigators within the ADNI contributed to the design and implementation of ADNI and/or provided data but did not participate in the analysis or writing of this report. A complete listing of ADNI investigators can be found at: https://adni.loni.usc.edu/wp-content/uploads/how_to_apply/ADNI_Acknowledgement_List.pdf

## Data Availability

The MRI and PET data from ADNI are available via the data access procedure described at http://adni.loni.usc.edu/data-samples/access-data/.

## Code Availability

The code developed for this work is available at

https://github.com/g-montana-valverde/GEC-TrophicLevels.

