## Supplementary Material for "Disrupted Brain Hierarchical Organization in Alzheimer’s Disease Progression"

### 1. ATN Biomarkers for Neuropathological Change

Consistent with previous research, Aβ distribution follows a spatial-temporal progression starting early in the neocortex and expanding to other brain areas as disease severity increases (Attems et al., 2005; Collij et al., 2021; Hampel et al., 2021) (Figure 2A). High Aβ accumulation (>50 centiloids; Maass et al., 2017) appear first in the insula, inferior temporal, orbitofrontal, fusiform, medial lobe (precuneus, cingulate, and dorsomedial prefrontal cortex), and amygdala as seen in HC+, then extend to frontal (superior and inferior) and paracentral areas in MCI+, and finally reaching parahippocampal, parietal and the visual cortices (cuneus, pericalcarine and lingual) in AD+. Group comparisons show a significant increase in Aβ accumulation across cortical networks.

In line with previous findings, abnormal tau accumulation begins in the entorhinal cortex and the hippocampus before spreading to neocortical areas in advanced AD+, parallel to the clinical worsening of the disease (Hof et al., 1991; Guillozet et al., 2003; Nelson et al., 2012; Figure 2B). High tau deposition (SUVR>1.3, Maass et al., 2017) is first detected in the middle and inferior temporal cortex, insula, precuneus, cingulate, putamen, amygdala, and hippocampus (HC- and HC+), before spreading to parietal and frontal regions in MCI+ and AD+. Tau accumulation is significantly higher in AD+ than in HC-, HC+, and MCI+, particularly within the VN, DAN, LN, CN, and DMN, with no differences in SMN or DAN.

Neurodegeneration, detected by GMV loss across brain regions and networks, follows the progression of the disease (Figure 2C). GMV progressively decreases in AD, beginning in the hippocampus and temporal lobes before the cingulate, precuneus, and frontal and parietal regions. This loss of volume is driven by neuronal loss and is closely associated with the decline in cognitive function seen in AD patients (Guo, et al., 2014; Wu et al., 2021). Network-level analyses reveal significant GMV reductions in the LN and DMN in AD+ relative to HC- and HC+, and in the DMN in MCI+ compared with HC+.

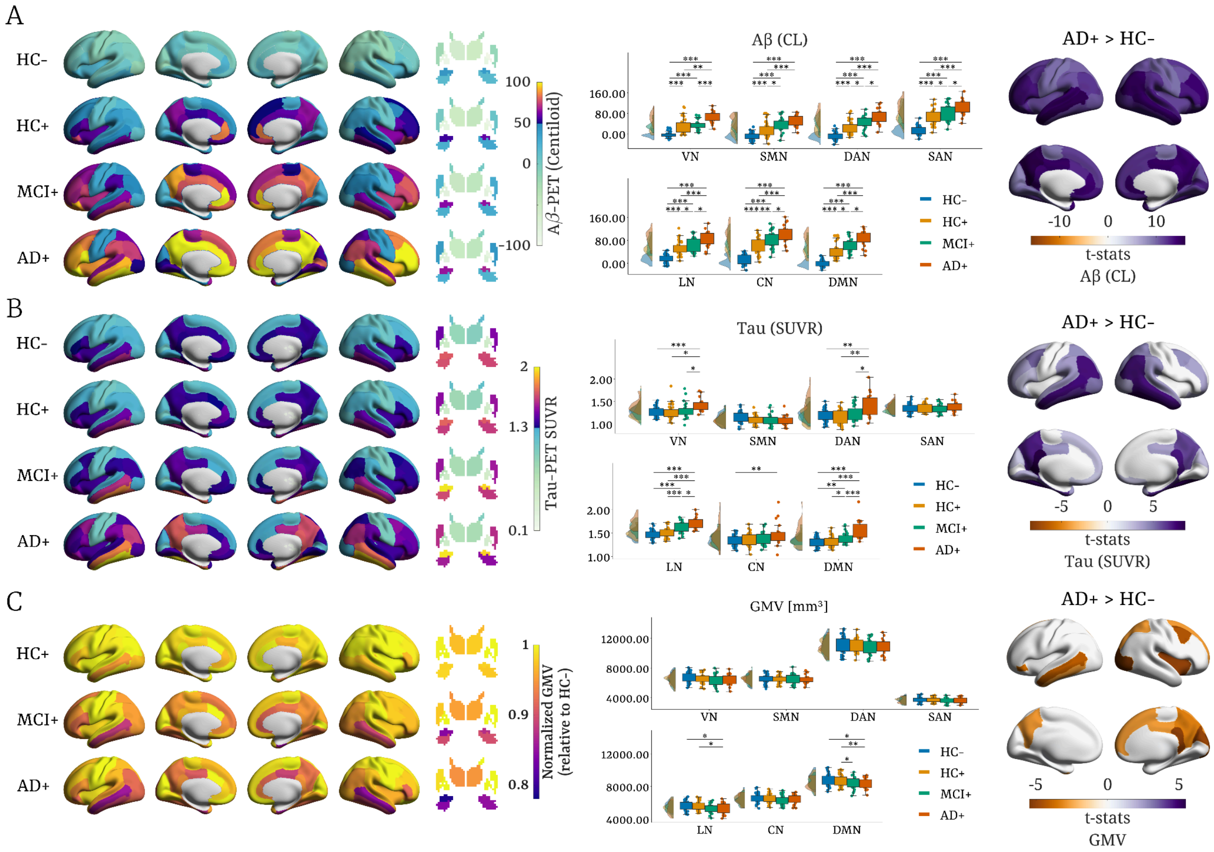


**Supplementary Figure 1. ATN biomarkers.** Cortical and subcortical representation of group average (A) Aβ and (B) Tau accumulation are shown in the brain renderings. Corresponding accumulations are averaged across functional networks in the boxplots aside. Additionally, region-level comparisons between AD+ and HC- are presented. (C) Normalized grey matter volume (GMV) relative to HC- is displayed in the renderings, with original GMV averaged across regions and region-level comparisons between AD+ and HC-. Group comparisons were performed using non-parametric permutation tests (10,000 permutations), controlling for age, gender, and education as covariates, and FDR-corrected. HC-: amyloid-negative healthy controls, HC+: amyloid-positive healthy controls, MCI+: amyloid-positive with mild cognitive impairment (MCI+), AD+: amyloid-positive with Alzheimer's disease. VN: visual network, SMN: somatomotor network, DAN: dorsal attention network, SAN: salience network, LN: limbic network, CN: control network, DMN: default mode network.

### 2. Changes in Cortical Trophic Levels During Task Performance

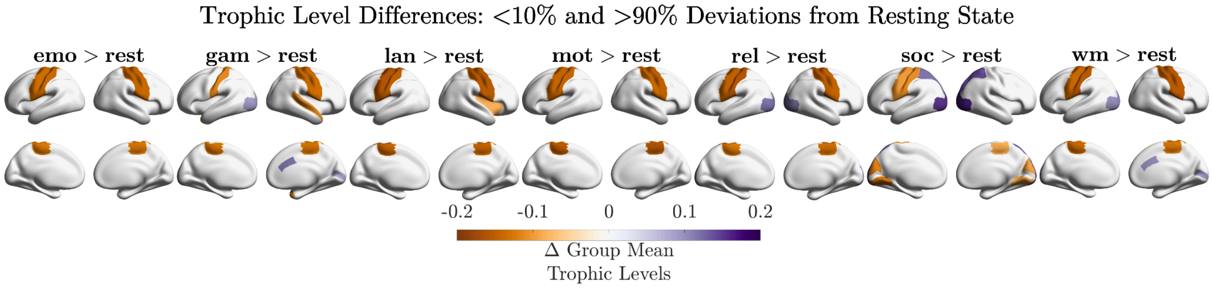


**Supplementary Figure 2. Lowest and highest significant trophic level deviations from resting-state.** Motor areas showed the greatest decrease in hierarchical position during task engagement, while visual areas exhibited the largest increase across several tasks, including gambling, relational, social and working memory. Statistical differences were computed by paired-permutation tests with FDR for multiple comparisons. emo: emotion; gam: gambling; lan: language; mot: motor; rel: relational; soc: social; wm: working memory.

### 3. Replication of Trophic Level Patterns in the Healthy Elderly Population

In order to assess the validation of the trophic level patterns in HC- individuals, we performed the same analysis on a comparable dataset from the Human Connectome Project in Aging (HCP-Aging, https://www.humanconnectome.org/study/hcp-lifespan-aging). This dataset collects brain imaging and behavioural data from adults aged 36-100+ for the understanding of how brain structure and function change with age. We selected a subset of participants with a matching mean age of 71.43 ± 2.59 years and healthy cognitive function as indicated by a MoCA total score greater than 26. This resulted in a final sample of 74 healthy elderly subjects.

From Supplementary Figure 3, we can infer that the hierarchical organization remains similar to HC- showing this gradient from sensorimotor and salience to visual and limbic networks. Positive correlation with imbalance ($r = 0.91$, $p < 0.001$) and negative correlation with mean path length ($r = -0.11$, $p < 0.001$) remain close to what we have measured in HC-, where no significant correlations were obtained with betweenness centrality and weakly significant correlations with clustering coefficient as well. These variables explained a substantial proportion of variance in trophic levels by an overall $R^2$ of 0.71. In addition, the correlation between directedness and the Frobenius norm of each individual's GEC remains significantly strong ($r = 0.899$, $p < 0.001$). Slight differences from the original dataset may arise from differences in scanner manufacturer, software version, and calibration procedures, in addition to differences in acquisition parameters. Notably, functional data was acquired with a TR of 0.8 s, while the original dataset used a TR=3 s.

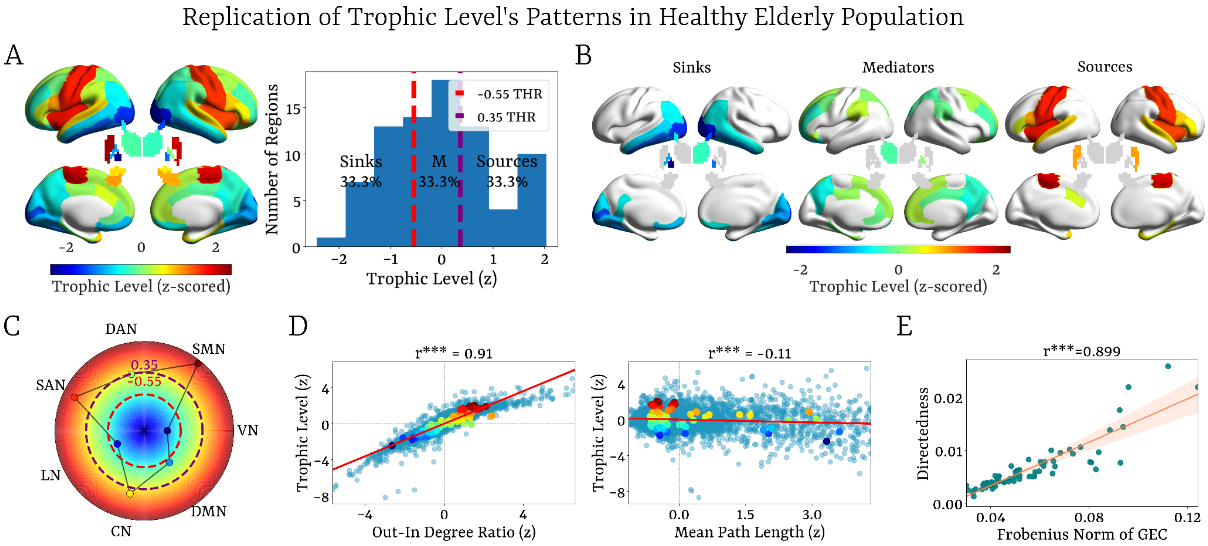


**Supplementary Figure 3. Trophic level's validation pattern in healthy elderly population from the HCP-Aging dataset. (A)** Z-scored trophic levels across the cortex and subcortex and their distribution were divided into three equal ranges, revealing the classification of brain regions into sinks, mediators, and sources. **(B)** Brain renderings displaying the three identified trophic level regimes (sinks, mediators, and sources) based on z-scored trophic levels. **(C)** Network level averaged trophic levels with corresponding thresholds for the distribution of trophic levels across the regions. **(D)** Correlations between z-scored trophic levels with the strongest and significant network properties, including the out-in-degree ratio ($r=0.90$, $p<0.001$) and mean path length ($r=-0.11$, $p<0.001$), showing the influence of causal imbalance and communication efficiency on trophic levels.

### 4. Machine Learning: ROC and SHAP

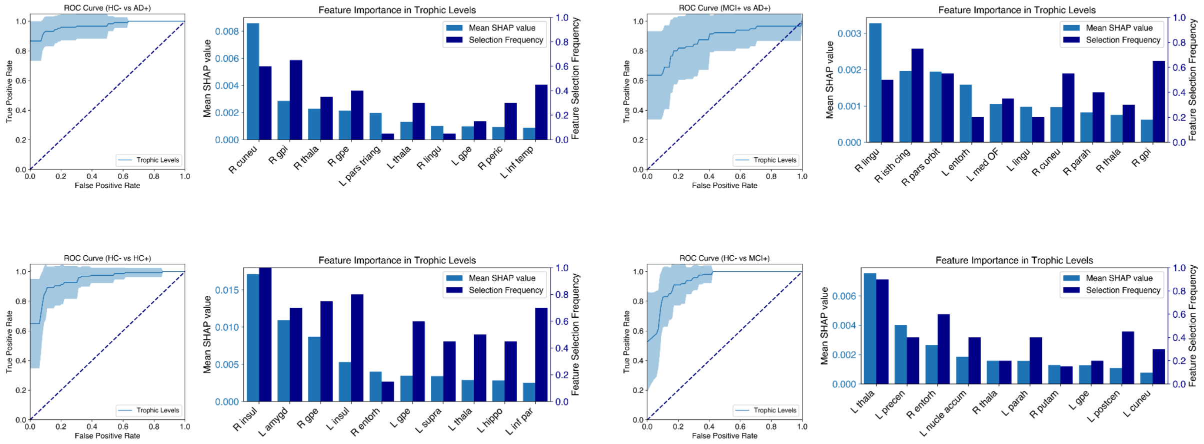


**Supplementary Figure 4.** Receiver Operating Characteristic (ROC) curves along with the highest SHapley Additive exPlanations (SHAP) feature importance values for the following group comparisons (from left to right and top to bottom): HC- vs AD+, MCI+ vs AD+, HC- vs HC+, and HC- vs MCI+.

### 5. Clinical Assessments Information

The MMSE (Folstein et al., 1975) assesses multiple cognitive domains, including orientation (time and place), registration (immediate word repetition), attention and concentration (serially subtracting seven beginning with 100), recall (recalling the previously repeated three words), language (naming, repetition, reading, writing, comprehension), and visual construction (copy two intersecting pentagons). Total scores range from 0 to 30, with lower values indicating poorer performance.

The CDR (Berg, 1989) provides a global rate of dementia severity based on the performance of cognitive functioning, including memory, orientation, judgment and problem solving, community affairs, home and hobbies, and personal care, synthesized into one global rating of dementia (ranging from 0 to 3). In this study, we use the CDR Sum of Boxes (CDR-SB), a more sensitive measure obtained by summing the scores across six domains (range: 0-18), with higher values indicating greater impairment. This provides a more detailed measure of cognitive and functional abilities compared to the single global score CDR.

The ADAS-Cog-13 (Rosen et al., 1984) is a structured, examiner-administered scale designed to assess memory, reasoning, language, orientation, and ideational and constructional praxis. It also evaluates spoken language, language comprehension, word-finding difficulties, and the ability to remember test instructions. Higher scores reflect worse cognitive performance.

The MoCA (Nasreddine et al., 2005) is a cognitive screening tool, similar to the MMSE, but specifically designed to detect participants at the MCI stage. It assesses multiple cognitive domains, including attention, executive functions, memory, language, visuospatial skills, conceptual thinking, calculations, and orientation. The test has a maximum of 30 points, with higher scores reflecting better cognitive performance.

For each participant, we included the cognitive assessments corresponding to the same study phase as the MRI data (ADNI3) and available for the same individuals in the demographics dataset. Only baseline or screening visits (e.g., *bl*, *sc*) were kept, ensuring that the cognitive tests were conducted closest to the corresponding imaging session. In cases where there were multiple tests for the same participant, the earliest visit was selected. Further details in clinical assessment protocols can be found in ADNI (https://adni.loni.usc.edu/help-faqs/adni-documentation/).

### 6. Image Pre-processing with *fMRIPrep*:

Results included in this manuscript come from preprocessing performed using *fMRIPrep* 21.0.1 (Esteban, Markiewicz, et al., 2019; Esteban, Blair, et al., 2018), which is based on *Nipype* 1.6.1 (Gorgolewski et al., 2011).

The T1-weighted (T1w) image was corrected for intensity non-uniformity (INU) with *N4BiasFieldCorrection* (Tustison et al., 2010), distributed with ANTs 2.3.3 (Avants et al., 2008), and used as T1w-reference throughout the workflow for each participant. The T1w-reference was then skull-stripped with a *Nipype* implementation of the *antsBrainExtraction.sh* workflow (from ANTs), using OASIS30ANTs as the target template. Brain tissue segmentation of cerebrospinal fluid (CSF), white-matter (WM) and grey-matter (GM) was performed on the brain-extracted T1w using *fast* (FSL 6.0.5.1:57b01774) (Zhang et al., 2002). Brain surfaces were reconstructed using *recon-all* (FreeSurfer 6.0.1) (Dale et al., 1999), and the brain mask estimated previously was refined with a custom variation of the method to reconcile ANTs-derived and FreeSurfer-derived segmentations of the cortical grey matter of Mindboggle (Klein et al., 2017). Volume-based spatial normalisation to one standard space (MNI152NLin2009cAsym) was performed through nonlinear registration with *antsRegistration* (ANTs 2.3.3), using brain-extracted versions of both T1w reference and the T1w template.

For each of the BOLD runs per subject, the following preprocessing was performed: First, a reference volume and its skull-stripped version were generated using a custom methodology of fMRIPrep. Head-motion parameters with respect to the BOLD reference (transformation matrices and six corresponding rotation and translation parameters) are estimated before any spatiotemporal filtering using *mcflirt* (FSL 6.0.5.1:57b01774) (Jenkinson et al., 2002). BOLD runs were slice-time corrected to 1.46s (0.5 of the slice acquisition range 0s-2.92s) using 3dTshift from AFNI (Cox & Hyde, 1997). The BOLD time-series (including slice-timing correction when applied) were resampled onto their original, native space by applying the transforms to correct for head-motion. These resampled BOLD time-series will be referred to as *preprocessed BOLD in original space*, or just *preprocessed BOLD*. The BOLD reference was then co-registered to the T1w reference using *bbregister* (FreeSurfer), which implements boundary-based registration (Greve & Fischl, 2009). Co-registration was configured with six degrees of freedom. Several confounding time-series were calculated based on the *preprocessed BOLD*: framewise displacement (FD), DVARS, and three region-wise global signals. FD was computed using two formulations following Power (absolute sum of relative motions) (Power et al., 2014) and Jenkinson (relative root mean square displacement between affines) (Jenkinson et al., 2002). FD and DVARS are calculated for each functional run, both using their implementations in *Nipype* (following the definitions by Power et al., 2014). The three global signals were extracted within the CSF, the WM, and the whole-brain masks. Additionally, a set of physiological regressors was extracted to allow for component-based noise correction (*CompCor*) (Behzadi et al., 2007). Principal components were estimated after high-pass filtering the *preprocessed BOLD* time-series (using a discrete cosine filter with 128s cut-off) for the two *CompCor* variants: temporal (tCompCor) and anatomical (aCompCor). tCompCor components are then calculated from the top 2% variable voxels within the brain mask. For aCompCor, three probabilistic masks (CSF, WM, and combined CSF+WM) are generated in anatomical space. The implementation differs from that of Behzadi et al. (2007) in that, instead of eroding the masks by 2 pixels on BOLD space, the aCompCor masks are subtracted from a mask of pixels that likely contain a volume fraction of GM. This mask is obtained by dilating a GM mask extracted from the FreeSurfer's *aseg* segmentation, and it ensures components are not extracted from voxels containing a minimal fraction of GM. Finally, these masks are resampled into BOLD space and binarized by thresholding at 0.99 (as in the original implementation). Components are also calculated separately within the WM and CSF masks. For each CompCor decomposition, the *k* components with the largest singular values are retained, such that the retained components' time series are sufficient to explain 50 percent of variance across the nuisance mask (CSF, WM, combined, or temporal). The remaining components are dropped from consideration. The head-motion estimates calculated in the correction step were also placed within the corresponding confounds file. The confound time series derived from head motion estimates and global signals were expanded with the inclusion of temporal derivatives and quadratic terms for each (Satterthwaite et al., 2013). Frames that exceeded a threshold of 0.5 mm FD or 1.5 standardised DVARS were annotated as motion outliers. All resamplings can be performed with *a single interpolation step* by composing all the pertinent transformations (i.e. head-motion transform matrices, susceptibility distortion correction when available, and co-registrations to anatomical and output spaces). Gridded (volumetric) resamplings were performed using *antsApplyTransforms* (ANTs), configured with Lanczos interpolation to minimise the smoothing effects of other kernels (Lanczos, 1964). Non-gridded (surface) resamplings were performed using *mri_vol2surf* (FreeSurfer).

Many internal operations of *fMRIPrep* use *Nilearn* 0.8.1 (Abraham et al., 2014), mostly within the functional processing workflow. For more details, see the fMRIprep website (https://fmriprep.org).

### 7. Derivation of Structural Connectomes from Diffusion MRI

Diffusion-weighted images (DWI) were preprocessed using MRTrix3 and FSL functions (Tournier et al., 2019; Smith et al., 2004). First, DWI were denoised using *dwidenoise* (Veraart et al., 2016), before preprocessing, applying FSL TOPT and EDDY to correct susceptibility distortions (Andersson et al., 2003) and eddy currents (Andersson & Sotiropoulos, 2016) using *dwifslpreproc*. Then, a B1 bios-filed correction was applied using *dwibiascorrect* (ANTs) (Tustison et al., 2010), followed by whole-brain mask generation (*dwi2mask*). The multi-tissue Dhollander method (*dwi2response*, Dhollander et al., 2016) was then used to determine tissue-specific response functions for white matter, grey matter, and CSF, to later obtain fibre orientation distributions (FODs) using spherical deconvolution (*dwi2fod,* Jeurissen et al., 2014), before normalization with *mtnormalise*. The T1-weighted anatomical images and 5-tissue segmentation were registered to the diffusion space using the *flirt* function to derive the grey-white matter interface for anatomically constrained tractography (ACT; Smith et al., 2012).

Finally, whole-brain probabilistic tractography was performed with ACT and backtracking (*tckgen*) by seeding at the GM/WM boundary, and generating 5 million streamlines per participant (maximum length 250 mm, FOD cutoff 0.06). The tractograms were then filtered using SIFT2 (*tcksift2*) to improve the quantitative correspondence between streamline densities and underlying FODs. Each participant's DK80 atlas was registered to diffusion space with FSL FLIRT using nearest-neighbour interpolation, and structural connectivity matrices were constructed with *tck2connectome*, using symmetric matrices with zero diagonal and SIFT2 weights. The resulting connectomes represent the streamline-weighted structural connectivity between the 80 parcellated brain regions.

### 8. Harmonization Tests ComBat

#### Grey Matter Volume Degeneration with Age - Healthy Controls

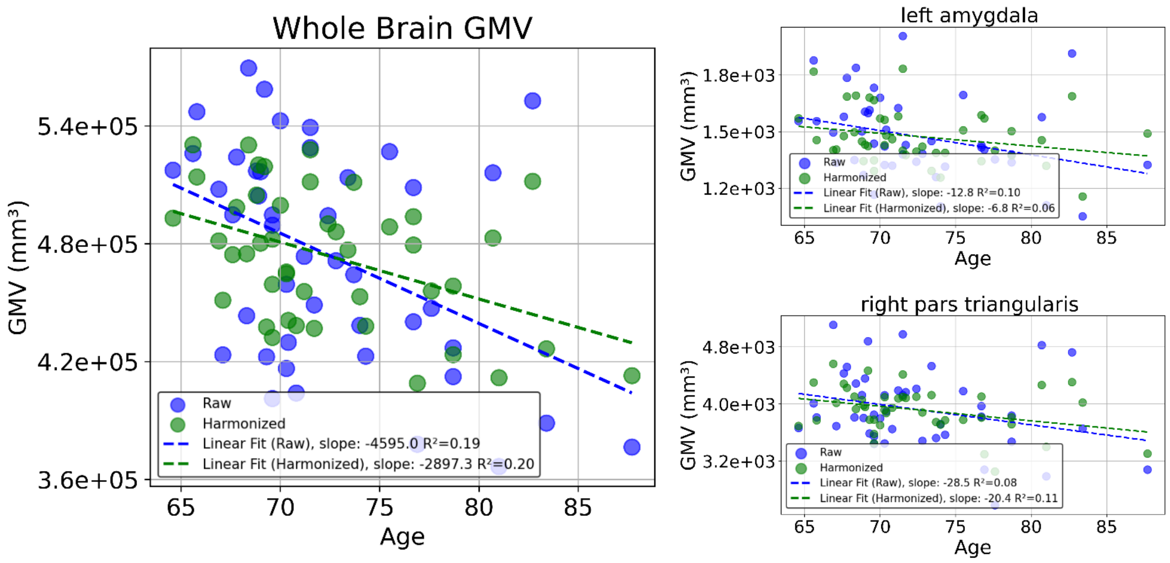


**Supplementary Figure 5. Grey Matter Volume (GMV) Degeneration with Age Before and After ComBat-Harmonization in Amyloid-Negative Healthy Controls (HC-), Simple Linear Regressions.** Whole Brain GMV, along with two examples, left amygdala and right pars triangularis, shows a significant decrease in GMV with age that remains after harmonization, establishing a ground-truth biological effect that is preserved while site-related variability is removed.

#### Trophic Levels Within-Group Differences

A

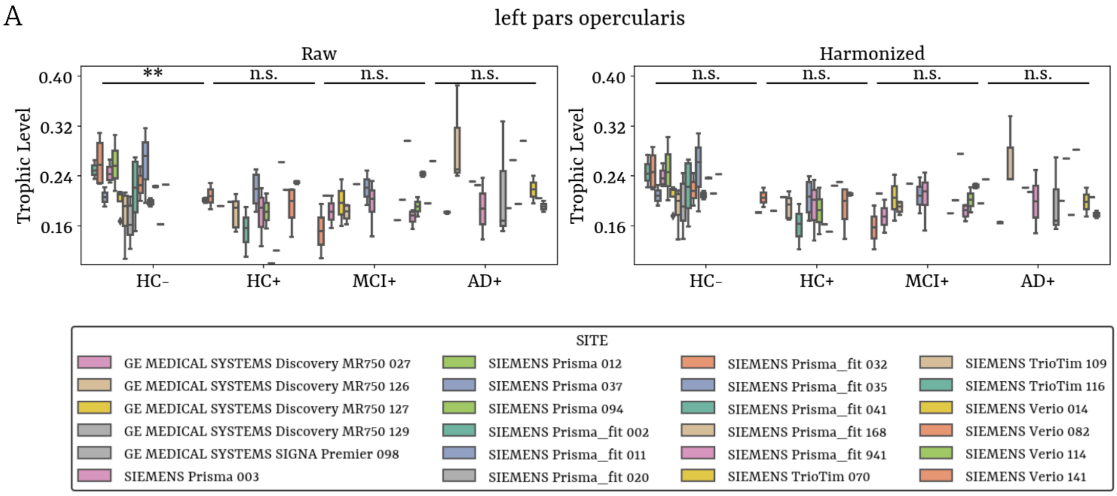


B

| | HC- | | HC+ | | MCI+ | | AD+ | |
|---|---|---|---|---|---|---|---|---|
| | HC- (raw) | HC- (harm.) | HC+ (raw) | HC+ (harm.) | MCI+ (raw) | MCI+ (harm.) | AD+ (raw) | AD+ (harm.) |
| left pars opercularis | ** | n.s. | n.s. | n.s. | n.s. | n.s. | n.s. | n.s. |
| left insula | n.s. | n.s. | n.s. | n.s. | * | n.s. | n.s. | n.s. |
| left amygdala | * | n.s. | n.s. | n.s. | n.s. | n.s. | n.s. | n.s. |
| right caudate | ** | n.s. | n.s. | n.s. | n.s. | n.s. | n.s. | n.s. |
| right medial orbitofrontal | * | n.s. | n.s. | n.s. | n.s. | n.s. | n.s. | n.s. |
| right entorhinal | n.s. | n.s. | n.s. | n.s. | * | n.s. | n.s. | n.s. |

**Supplementary Figure 6. Within-Group Trophic Level Differences Before and After ComBat-Harmonization in Amyloid-Negative Healthy Controls (HC-), Amyloid-Positive Healthy Controls (HC+), Amyloid-Positive individuals with Mild Cognitive Impairment (MCI+), and Amyloid-Positive patients with Alzheimer's Disease (AD+). (A)** Example from the left pars opercularis showing significant within-group differences in HC- which vanish after harmonization. **(B)** Table listing all regions that initially showed significant differences, all of which lost significance after harmonization. Within-group differences across sites were assessed using ANOVA.

### 9. Generative Effective Connectivity (GEC) Methodology

#### The Hopf Model

To model the whole-brain dynamics requires modelling the coupling of the local dynamics of $N$ Hopf nodes interconnected through a given coupling matrix, i.e., by adding a diffusive coupling term representing the input received in node $j$ from every other node $i$, which is weighted by the corresponding general effective connectivity (GEC), $C_{ji}$. This input is modelled using the common difference coupling, which approximates the simplest (linear) part of a general coupling function (Kringelbach et al., 2023). Then, the stochastic coupled nonlinear differential equation, i.e., the Stuart-Landau oscillators, gives the state variables:

$$\frac{dz_j}{dt} = (a_j + i\omega) z_j - |z_j|^2 z_j + \sum_{i=1}^{N} G_{ji} (z_i - z_j) + \eta_j \tag{1}$$

where the complex variable (which has arbitrary units)

$$z_j = x_j + iy_j \tag{2}$$

denotes the temporal evolution of the activity in node $j$; $|z_j|$ is the module of $z_j$, i.e., $|z_j|^2=x_j^2+y_j^2$; $\omega=2\pi\upsilon$ is the intrinsic angular frequency (in *rad/s*), where $\upsilon$ is the intrinsic frequency in Hz; the parameter $a_j$ is the local bifurcation parameter (in $s^{-1}$) and constant across nodes (i.e., $a_j = a$); finally, η is the additive white noise, i.e., $\langle\eta(t)\rangle = 0$ and $\langle\eta(t)\eta(t')\rangle = \sigma^2\delta(t-t')$, where σ is the noise amplitude (in $s^{-1/2}$) fixed to $\sigma^2=0.01$ (Kringelbach et al., 2023), and the angular brackets $\langle.\rangle$ denote the average over stochastic realisations. The intrinsic frequencies $\upsilon$ were estimated from empirical data as the averaged peak frequencies of the narrowband BOLD signals of the different nodes (in a 0.008 - 0.08 Hz band, as in Kringelbach et al. 2023).

For $a_j < 0$, the fluctuations of the local dynamics are attenuated, indicating that the system relaxes, presenting a stable spiral point producing damped or noisy oscillations in the absence or presence of noise, respectively, in a so-called subcritical regime. On the other hand, if $a_j > 0$, the system produces self-sustained oscillations, i.e., a stable limit cycle, with a constant amplitude and constant intrinsic frequency $\omega_j/(2\pi)$, in a so-called supercritical regime (Supplementary Figure 7).

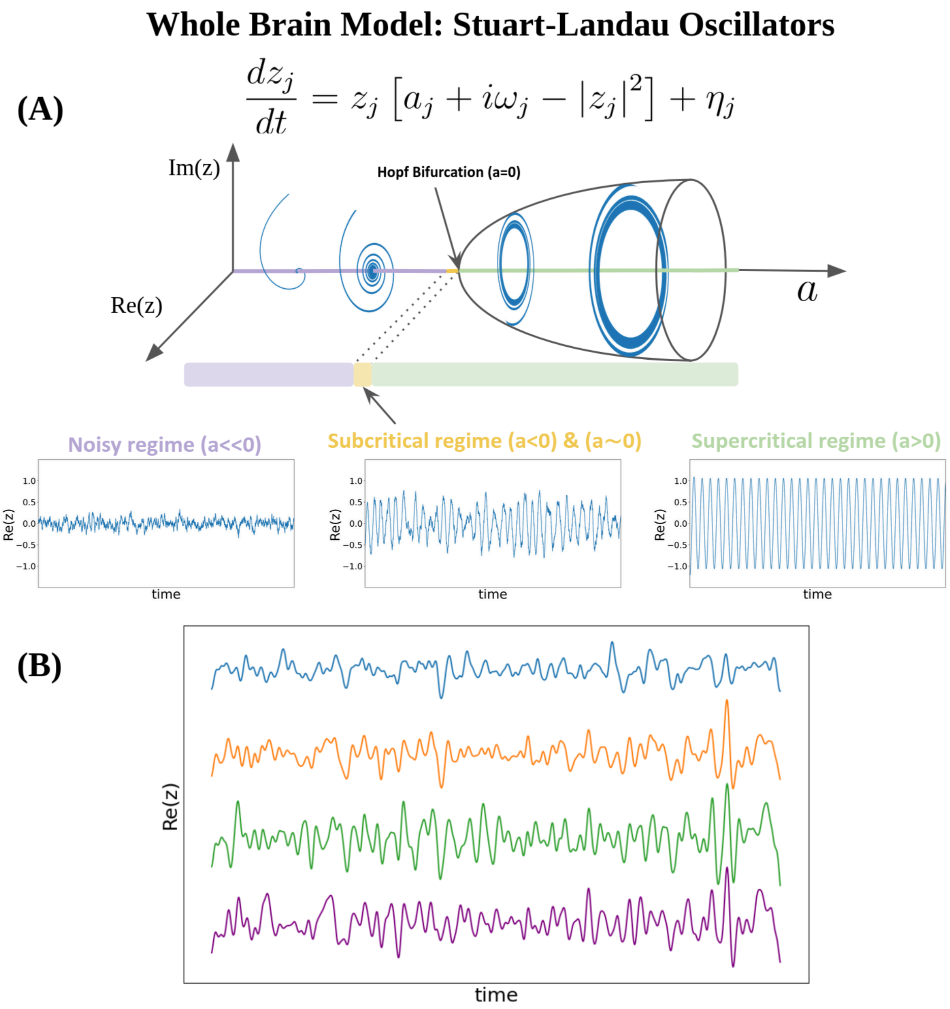


**Supplementary Figure 7**. **(A)** The Stuart-Landau Oscillator produces three different signals given a local bifurcation parameter *a*: 1) Noisy regime - a noise signal resulting from Gaussian noise when the parameter is much less than zero; 2) Subcritical regime - a fluctuating stochastically structured signal when the parameter is just below zero, which allows the system to fluctuate between noise and oscillations; 3) Supercritical regime - an oscillatory signal when the parameter is larger than zero. Parameters: $a_j = -0.5, \ -0.1, \ 1$; $\omega_j = 5\, rad\cdot s^{-1}$ and $\sigma = 0.3$. **(B)** Brain signals are modelled by the real part of the state variables, i.e., *x=Re(x)*. Illustrated are four different brain BOLD signals from the Human Connectome Project (HCP) using Schaefer parcellation with N=100 nodes.

### Linearization of the Hopf Model

Because the brain operates near criticality (Chialvo, 2010; Tagliazucchi et al., 2012; Haimovici et al., 2013; Tagliazucchi et al., 2017), the best working point for fitting whole-brain dynamics is at the brink of the Hopf bifurcation, i.e., for the bifurcation parameter *a* at the edge of zero on the negative side, such that the oscillators remain damped still. Previous findings have demonstrated that at this region the correlation between the empirical and the simulated FC is maximised, more specifically, at $a_j = -0.02$ (Deco et al., 2017; Sanz Perl et al., 2022; Kringelbach et al., 2023).

Previous whole-brain model comparisons showed that non-linearities are minimal. This finding enables us to avoid the nonlinear coupling, thereby simplifying the network statistics without requiring extensive numerical simulations. This suggests that the brain operates in the simpler noisy-oscillation regime, and one can estimate the statistics of the whole brain network (e.g., variances and covariances) using a linear approximation (Piccinini et al., 2022; Ponce-Alvarez & Deco, 2024).

The functional correlations between all pairs of brain regions can be estimated by employing a linear noise approximation (LNA), so equation (1) can be rewritten as:

$$\frac{d\mathbf{z}}{dt} = (\mathbf{a} - \mathbf{S} + i\boldsymbol{\omega}) \odot \mathbf{z} - (\mathbf{z} \odot \bar{\mathbf{z}})\,\mathbf{z} + \mathbf{G}\mathbf{z} + \boldsymbol{\eta} \tag{1s}$$

where $\mathbf{z} = [z_1, ..., z_N]^T$ , $\mathbf{a} = [a_1, ..., a_N]^T$, $\boldsymbol{\omega} = [\omega_1, ..., \omega_N]^T$, $\boldsymbol{\eta} = [\eta_1, ..., \eta_N]^T$ represents the vector of uncorrelated noise, and $\mathbf{S} = [S_1, ..., S_N]^T$ is the vector containing the connectivity strength of each node, i.e, $S_i = \Sigma_j C_{ij}$ . The superscript *T* represents the transpose, $\odot$ the Hadamard element-wise product, i.e., $u \odot v = [u_1 v_1, ..., u_N v_N]^T$, and $\bar{\mathbf{z}}$ the complex conjugate of $\mathbf{z}$. This equation represents the linear fluctuations $\delta\mathbf{z}$ around the fixed point $\mathbf{z} = 0$, which is the solution to the equation $\frac{d\mathbf{z}}{dt} = 0$. Separating the real and imaginary parts and discarding the higher-order terms $(\mathbf{z} \odot \bar{\mathbf{z}})\,\mathbf{z}$, the evolution of the linear fluctuations $\delta\mathbf{u}$ follows a Langevin stochastic linear equation:

$$\frac{d}{dt}\delta\mathbf{u} = \mathbf{J}\delta\mathbf{u} + \boldsymbol{\eta}\,, \tag{2s}$$

where $\delta\mathbf{u} = [\delta\mathbf{x}, \delta\mathbf{y}]^T = [\delta x_1, \cdots, \delta x_N, \delta y_1, \cdots, \delta y_N]^T$ is a 2N-dimensional column vector that contains the fluctuations of real and imaginary state variables. The matrix $\mathbf{J}$ corresponds to the Jacobian of the system evaluated at the fixed point, which can be written as a $2N \times 2N$ matrix

$$\mathbf{J} = \begin{bmatrix} \mathbf{J_{xx}} & \mathbf{J_{xy}} \\ \mathbf{J_{yx}} & \mathbf{J_{yy}} \end{bmatrix}, \tag{3s}$$

where $\mathbf{J_{xx}}$, $\mathbf{J_{yy}}$, $\mathbf{J_{xy}}$, $\mathbf{J_{yx}}$, are $N \times N$ matrices, $\mathbf{J_{xx}} = \mathbf{J_{yy}} = diag\,(\mathbf{a} - \mathbf{S}) + \mathbf{G}$, and $\mathbf{J_{xy}} = -\mathbf{J_{yx}} = diag(\boldsymbol{\omega})$, a diagonal matrix whose diagonal is a vector $\boldsymbol{\omega}$. The above linearization is only valid if $\mathbf{z} = 0$ is a stable solution of the system. Note that all eigenvalues of the Jacobian have a negative real part.

To model the functional connectivity, we must first compute the covariance of the fluctuations around the origin, i.e., $\mathbf{K} = \langle \delta\mathbf{u}\delta\mathbf{u}^T \rangle$ describing how the fluctuations between regions are related to each other. We begin by writing equation (2s) as $d\delta\mathbf{u} = \mathbf{J}\delta\mathbf{u}dt + d\mathbf{W}$, where $\mathbf{W}$ is a $2N - dimensional$ Wiener process (Brownian motion) with covariance $\langle d\mathbf{W}d\mathbf{W}^T \rangle = \mathbf{Q_n}dt$ , and $\mathbf{Q_n} = \langle \boldsymbol{\eta}\boldsymbol{\eta}^T \rangle$ is the covariance matrix of the noise ($\mathbf{Q_n}$ is diagonal if the noise is uncorrelated, i.e., $\mathbf{Q_n} = \sigma^2\mathbf{I}$). Then, using Itô's stochastic calculus, we have $d(\delta\mathbf{u}\delta\mathbf{u}^T) = d\,(\delta\mathbf{u})\,\delta\mathbf{u}^T + d\delta\mathbf{u}\,(\delta\mathbf{u}^T) + d\,(\delta\mathbf{u})\,d\,(\delta\mathbf{u}^T)$. Since $\langle \delta\mathbf{u}d\mathbf{W}^T \rangle = 0$ and keeping terms in first order in the differential $dt$ (as $dt^2$ can be made arbitrarily small), we obtain $d\,\langle \delta\mathbf{u}\delta\mathbf{u}^T \rangle = \mathbf{J}\,\langle \delta\mathbf{u}\delta\mathbf{u}^T \rangle\,dt + \langle \delta\mathbf{u}\delta\mathbf{u}^T \rangle\,\mathbf{J}^T dt + \mathbf{Q_n}dt$, and so:

$$\frac{d\mathbf{K}}{dt} = \mathbf{JK} + \mathbf{KJ^T} + \mathbf{Q}. \tag{4s}$$

Hence, the stationary covariances, for which $\frac{d\mathbf{K}}{dt} = 0$, can be obtained solving the analytic Lyapunov equation using the eigen-decomposition of the Jacobian matrix,

$$\mathbf{JK} + \mathbf{KJ^T} + \mathbf{Q} = 0. \tag{5s}$$

We then obtain the simulated functional connectivity $FC^{model}$ from the first $N$ rows and columns of the covariance $\mathbf{K}$, which corresponds to the real part of the dynamics, precisely, representing the BOLD fMRI signal.

### 10. Hierarchy Metrics: Trophic Levels

Here, the directed graph is represented by $G(V, W, \Delta)$, where $V = \{1, \ldots, n\}$ is a set of $n$ nodes, $W$ is the $n \times n$ matrix of weights, and $\Delta$ is a $n \times n$ matrix of $\delta_{ij}$ elements that represent the desired hierarchical difference along the y-axis between nodes $i$ and $j$, based on the structure of the graph (i.e., which node should be placed above or below another). If $\delta_{ij} > 0$, node $i$ should be placed above node $j$, and if $\delta_{ij} < 0$, node $i$ should be placed below $j$. By definition, $\Delta$ is an antisymmetric matrix, $\delta_{ij} = -\delta_{ji}$. The weight associated with the edge connecting nodes $i$ and $j$ is written by $w_{ij} \geq 0 \quad \forall i, j$. Self-edges $(i, i)$, or loops, are permitted.

The conventional definition of the Laplacian for non-symmetric weighted directed graphs is the $n \times n$ matrix

$$L_{ij} = \begin{cases} \sum_{k=1}^{n} (w_{ik} + w_{ki}) & if\ i = j, \\ -w_{ij} - w_{ji} & if\ i \neq j \end{cases} \quad for \quad i, j = 1, 2, \ldots, n. \tag{6s}$$

The balance of the $i'th$ node measures the difference between how much influence it exerts on other nodes (those nodes $j$ for which $\delta_{ij} > 0$), and how much it is influenced (those nodes $j$ for which $\delta_{ij} < 0$):

$$b_i = \sum_{j=1}^{n} w_{ij} \delta_{ij}. \tag{7s}$$

For weighted directed graphs is simply the difference between the out-degree and the in-degree of each node; thus, the name balance: a node with a positive balance is more influential, whereas a negative balance indicates that it is more influenced.

The idea of using energy minimization for layering was already exploited in the field of undirected graph representations. Two of the most successful realizations of this idea were introduced by Tutte (1963) and Hall (1970): the solution to the problem of placing $n$ connected nodes on a y-axis is obtained by minimizing the energy function, or hierarchy energy, which is equal to the weighted sum of the squared distances between the nodes:

$$E_H = \frac{1}{2} \sum_{i,j=1}^{n} w_{ij} (y_i - y_j)^2, \tag{8s}$$

where $y_i$ denotes the Y-coordinate of node $i$, $E_H$ the hierarchy energy, and the squared difference is introduced to avoid negative values inside the sum. Then, the problem for undirected graphs is to find the row vector $y = (y_1, \ldots, y_n)^T$ that corresponds to the minimal relative height positions $y_i - y_j$ for any pair of nodes. The idea is driven by reducing as its minimum the tension of the graph, encouraging connected nodes to have similar positions. The weight $w_{ij}$ represents the 'importance' of the connection, and the squared difference in positions, $(y_i - y_j)^2$, penalizes large separations.

The quadratic form of the equation guarantees a unique global minimum, the subtraction minimizes the deviation from the target hierarchy difference, taking into account the weight influences, and preservation of directionality, ensuring that each edge influences the relative position of each pair of nodes.

For directed weighted graphs, the energy function is adjusted to account for directionality,

$$E_H = \frac{1}{2}\sum_{i,j=1}^{n} w_{ij}\left(y_i - y_j - \delta_{ij}\right)^2 = \sum_{(i,j)\in E} w_{ij}\left(y_i - y_j - \delta_{ij}\right)^2, \tag{9s}$$

where the goal is to arrange the nodes to minimize the discrepancy between their actual differences $y_i - y_j$ and the desired differences $\delta_{ij}$. The weight $w_{ij}$ scales this discrepancy, indicating the importance of respecting the target height difference for that particular edge. In other words, the larger the weight, the smaller should be $\left(y_i - y_j - \delta_{ij}\right)^2$ to keep the contribution to the energy small.

The hierarchy energy can be written in a matrix form using the previously mentioned Laplacian and balance (equations 6s and 7s):

$$E_H(G,y) = E_0 + y^T L y - 2y^T b, \tag{10s}$$

where $E_0 = \frac{1}{2}\sum_{i,j=1}^{n} w_{ij}\delta_{ij}^2$.

The optimal arrangement of the graph that minimizes the hierarchy energy is $y^* = arg\min_y E_H(G,y)$, obtained by calculating the global minimum of equation 10s:

$$\left.\frac{\partial E_H}{\partial y}\right|_{y=y^*} = 2Ly^* - 2b = 0, \tag{11s}$$

that is the solution of the system

$$Ly^* = b. \tag{12s}$$

Note that the number of solution is infinite: when $G$ is a connected graph, the Laplacian, $L$, has exactly one zero eigenvalue, corresponding to the eigenvector $c \cdot 1_n$; in other words, the system is under-constrained in terms of absolute positions, specifically, translations of all node positions by the same constant do not affect the structure of the graph, so we can shift the entire solution $y^*$ by any constant $c$ maintaining the differences unchanged (Hall, 1970). Nevertheless, all these infinite solutions only differ in translation so, in the end, we only have a unique value of $y^*$ (Carmel et al., 2002).

### 11. Hierarchical Metrics Comparison: Reciprocity, Non-normality, Directedness

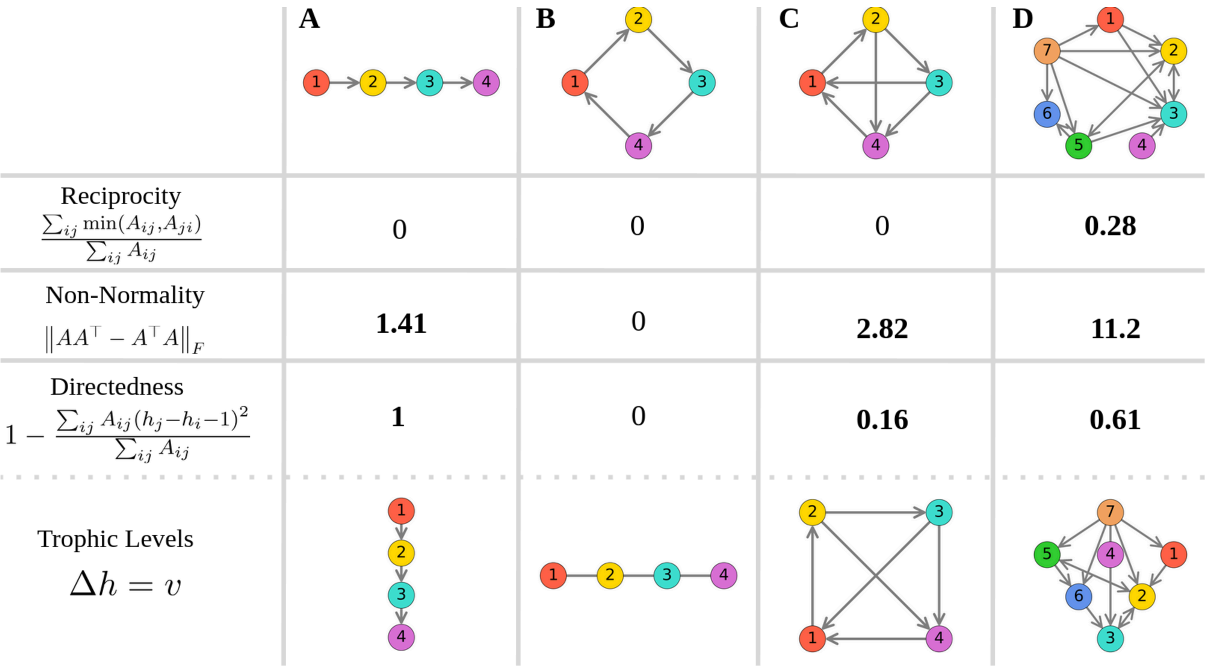


**Supplementary Figure 8. Trophic directedness is more sensitive to global patterns when exploring the hierarchical structure of a directed network.** This figure shows simple unidirected graph configurations. While reciprocity fails to differentiate between a chain **(A)**, a cycle **(B)**, and an interconnected graph **(C)**, non-normality also is not able to capture the hierarchical nature of the chain– something that is well revealed by the directedness measure.

### 12. Graph Metrics

For each individual's directed graph *C*, the out-in-degree ratio of each node *n* was computed from the in-weights ($d_n^{in}$) and the out-weights ($d_n^{out}$) given in the previous section:

$$r_i = \frac{d_n^{out}}{d_n^{in}},$$

for $d_n^{in} \neq 0$.

The weighted clustering coefficient $Clu_n$ was estimated following the Brain Connectivity Toolbox formulation for directed graphs (Fagiolo, 2007). Defining the symmetrized weight matrix raised to $1/3$, $s_{ij} = w_{ij}^{1/3} + w_{ji}^{1/3}$, the total degree

$k_i = \sum_j (a_{ij} + a_{ji})$ of the binary adjacency $a_{ij} = 1$ if $w_{ij} \neq 0$, else, $0$, we can compute the clustering coefficient as

$$Clu_i = \frac{cyc3_i}{CYC3_i} = \frac{1/2\,(S^3)_{ii}}{k_i\,(k_i - 1) - 2\,(A^2)_{ii}},$$

where $cyc3_i$ is the number of weighted directed triangles (3-cycles) and $CYC3_i$ the number of possible directed triangles in a node $i$.

The mean path length $L_i$ was obtained from all pairs of shortest paths using Dijkstra's algorithm, where the distance between two connected nodes is defined as the inverse of their connection weights, i.e., $d(m,n) = \frac{1}{w_{mn}}$ for $w_{mn} > 0$, else, inf if $w_{mn} = 0$. Then, for every node $i$, the shortest-path distance will be given by:

$$L_{ij} = \min_{paths\ i \to j} \sum_{(m,n) \in path} d(m,n)\ .$$

Finally, for each node $i$, the mean path length is the mean distance to all reachable nodes:

$$MeanPathLength(i) = \frac{1}{|R_i|} \sum_{j \in R_i} L_{ij}\ ,$$

where $R_i$ is the set of nodes reachable from $i$.

The betweenness centrality $C_B$ of the node $i$ is:

$$C_B(i) = \sum_{s \neq v \neq t} \frac{\sigma\,(s, t|v)}{\sigma\,(s,t)}\ ,$$

where $\sigma\,(s,t)$ is the number of shortest paths from $s$ to $t$, and $\sigma\,(s,t|v)$ is the number of those shortest paths that pass through node $i$.
